# Mid-to-Late Life Healthy Lifestyle Modifies Genetic Risk for Longitudinal Cognitive Aging among Asymptomatic Individuals

**DOI:** 10.1101/2024.05.26.24307953

**Authors:** Yuexuan Xu, Zhongxuan Sun, Erin Jonaitis, Yuetiva Deming, Qiongshi Lu, Sterling C. Johnson, Corinne D. Engelman

## Abstract

**IMPORTANCE:** Genetic and lifestyle factors contribute to an individual’s risk of developing Alzheimer’s disease. However, it is unknown whether and how adherence to healthy lifestyles can mitigate the genetic risk of Alzheimer’s.

**OBJECTIVE:** The aim of this study is to investigate whether adherence to healthy lifestyles can modify the impact of genetic predisposition to Alzheimer’s disease on later-life cognitive decline.

**DESIGN, SETTING, AND PARTICIPANTS:** This prospective cohort study included 891 adults of European ancestry, aged 40 to 65, who were without dementia and had complete healthy-lifestyle and cognition data during the follow-up. Participants joined the Wisconsin Registry for Alzheimer’s Prevention (WRAP) beginning in 2001. We conducted replication analyses using a subsample with similar baseline age range from the Health and Retirement Study (HRS).

**EXPOSURES:** We assessed participants’ exposures using a continuous non-*APOE* polygenic risk score for Alzheimer’s, a binary indicator for *APOE-ε4* carrier status, and a weighted healthy-lifestyle score, including factors such as no current smoking, regular physical activity, healthy diet, light to moderate alcohol consumption, and frequent cognitive activities.

**MAIN OUTCOMES AND MEASURES:** We z-standardized cognitive scores for global (Preclinical Alzheimer’s Cognitive Composite score 3 – PACC3) and domain-specific assessments (delayed recall and immediate learning).

**RESULTS:** We followed 891 individuals for up to 10 years (mean [SD] baseline age, 58 [6] years, 31% male, 38% *APOE-ε4* carriers). After false discovery rate (FDR) correction, we found statistically significant PRS × lifestyle × age interactions on preclinical cognitive decline but the evidence is stronger among *APOE-ε4* carriers. Among *APOE-ε4* carriers, PRS-related differences in overall and memory-related domains between people scoring 0-1 and 4-5 regarding healthy lifestyles became evident around age 67 after FDR correction. These findings were robust across several sensitivity analyses and were replicated in the population-based HRS.

**CONCLUSION:** A favorable lifestyle can mitigate the genetic risk associated with current known non-*APOE* genetic variants for longitudinal cognitive decline, and these protective effects are particularly pronounced among *APOE-ε4* carriers.

**KEY POINTS:** *Question:* Can a healthy lifestyle modify genetic susceptibility to Alzheimer’s disease and its impact on cognitive decline?

*Findings:* In two longitudinal studies, we found that adhering to a healthy lifestyle can reduce the adverse genetic effects of known non-*APOE* variants, especially in *APOE-ε4* carriers. Specifically, *APOE-ε4* carriers with <2 healthy lifestyle factors had significantly higher genetic risk caused by currently known genetic variants compared to those with two or more.

*Meaning:* A healthy lifestyle can mitigate genetic risks from non-*APOE* variants, especially among *APOE-ε4* carriers. Future research should include biomarker analysis to uncover the underlying biological mechanisms.

## INTRODUCTION

The pathophysiological progression of late-onset Alzheimer’s disease (LOAD) occurs decades before the manifestation of clinical symptoms. Characterizing this pre-symptomatic period can facilitate the development of effective screening, prevention, and treatment strategies. Genetic and lifestyle factors affect an individual’s risk of developing LOAD. The *APOE* gene is the strongest genetic risk factor for LOAD, with the ε4 allele increasing the risk of LOAD and the ε2 allele conferring protection^1^. Recent genome-wide association studies have identified additional loci associated with the risk of LOAD even though most of them only exhibit miniscule effects^2–6^. The polygenic risk score (PRS), combining the effects of multiple independent risk alleles, is predictive of LOAD risk with up to 84% accuracy^7^.

A number of modifiable factors, smoking, physical activity, diet, cognitive activity, and alcohol consumption, may contribute individually to the risk of LOAD^8–12^. Recent studies have combined individual factors into a composite lifestyle score and demonstrated adherence to healthy lifestyle is associated with lower LOAD risk^13,14^. Although it has been hypothesized that a healthy lifestyle may mitigate the adverse genetic effect, existing studies have yielded mixed results^13,15–17^. In our earlier work, we found that the effects of *APOE* and non-*APOE* PRS are age-related, and the genetic burden of the non-*APOE* PRS is more detrimental among *APOE-ε4* carriers than among noncarriers^18,19^. These findings indicate that the risk modifying effect of nongenetic protective factors on non-*APOE* PRS may be best detected among *APOE-ε4* carriers.

The purpose of this study was to use data from two longitudinal studies to investigate whether following a healthy lifestyle can mitigate the genetic risk of LOAD on cognitive decline after considering the age heterogeneity and modifying effect of *APOE-ε4* carrier status on non-*APOE* PRS.

## METHODS

### Study settings and participants

Started in 2001, the Wisconsin Registry for Alzheimer’s Prevention (WRAP) is an ongoing prospective cohort study of >1700 late-middle-aged English-speaking adults who were non-demented at enrollment and enriched for a parental history of AD. Participants are followed approximately biennially until dementia, dropout, death, or the age of 85. At each visit, participants complete extensive neuropsychological testing and a physical examination, described elsewhere^20^. Participants eligible for the current study were those of European genetic ancestry. Visit 2 was considered as baseline for the current study because this was the first visit that included the cognitive composite scores of interest. Written informed consent was obtained from all participants before participation. The University of Wisconsin–Madison Institutional Review Board approved this study. Strengthening the Reporting of Observational Studies in Epidemiology (STROBE) reporting guidelines were followed. Data were collected between November 2006 and May 2022.

### Genotyping, APOE, and non-APOE PRS

WRAP DNA samples were genotyped using the Illumina Infinium Expanded Multi-Ethnic Genotyping Array (MEGA^EX^) at the University of Wisconsin Biotechnology Center. Details about genotyping and quality control have been described elsewhere^21^. The non-*APOE* PRS is a weighted sum (∼N(0,1)) of independent single-nucleotide polymorphisms that have shown genome-wide significant (*P*<5e^−8^, excluding *APOE*) association with AD ^2,22^. A higher PRS corresponds to a higher genetic risk of LOAD. *APOE* genotype was determined based on single-nucleotide polymorphisms, rs7412 and rs429358, and combined into two groups: individuals who were *APOE-ε4* carriers (ε2/ε4, ε3/ε4, ε4/ε4) and noncarriers (ε2/ε2, ε2/ε3, ε3/ε3).

### Healthy-lifestyle score

A healthy-lifestyle score was constructed using five well-established LOAD risk factors that are individually and collectively associated with risk of dementia (smoking status, physical activity, diet, alcohol consumption, and cognitive activity) based on self-reported lifestyle questionnaires^8–12^. Of the five indicators, smoking status, physical activity, and alcohol consumption were time varying. Because data on diet and cognitive activity were added to the WRAP study after baseline, we used the first visit at which these two measures were available to all participants (visit 5) and mapped visit 5 data to all other visits ^23^. We categorized each factor as healthy or unhealthy based on the guidelines provided by Dhana et al^13^. The criterion for each healthy behavior was: 1) not being a current smoker; 2) moderate or vigorous exercises for at least 150 minutes per week; 3) Mediterranean-DASH Diet Intervention for Neurodegenerative Delay (MIND) diet score (without alcohol) in the top 40% of the cohort distribution; 4) light to moderate alcohol consumption (1-7 drinks per week for women and 1-14 drinks per week for men); and 5) cognitive activities in the top 40% of the cohort distribution, as measured with the Florida Cognitive Activities Scale (FCAS). Currently, there is no threshold for healthy diet or cognitive activities to determine healthy/unhealthy behaviors; therefore, we imposed a 40% upper cutoff based on the cohort distribution, following the previous literature^13,24–26^. WRAP participants scored 1 for each of those factors if they met the “healthy” criterion and scored 0 otherwise. The final score for adherence to a healthy lifestyle ranged from 0 to 5, with a higher score indicating a healthier lifestyle.

### Cognition

In the current study, we assessed the participant’s global cognitive performance using Preclinical Alzheimer’s Cognitive Composite 3 (PACC-3), which is computed by standardizing and averaging performance from three tests, the Rey Auditory Verbal Learning (RAVLT; Trials 1–5), Wechsler Memory Scale-Revised (WMS-R) Logical Memory II total score (LMII), and Wechsler Adult Intelligence Scale-Revised (WAIS-R) Digit Symbol Coding, total items completed in 90 seconds^27,28^. We also assessed two domain-specific cognitive composites: immediate learning and delayed recall^29^. The immediate learning composite score was derived from the RAVLT total in trials 1–5, the WMS-R Logical Memory I total score (LMI), and the Brief Visuospatial Memory Test-Revised (BVMT-R) immediate-recall score. The delayed recall composite score was obtained from the sum of the RAVLT long-delay free recall score, the WMS-R logical memory delayed recall score, and the BVMT-R delayed recall score. A higher cognitive composite score indicates better cognitive performance.

### Statistical analyses

Our primary goal was to identify whether adherence to a healthy lifestyle can modify the effect of genetic predisposition to LOAD on later-life cognitive decline while accounting for the age-related genetic effect and the modifying effect of *APOE-ε4* carrier status on PRS. Using a linear mixed-effects model with random intercepts for subject and family, we first examined *APOE-ε4*×lifestyle×age and PRS×lifestyle×age interactions in the full sample. We then stratified the sample by *APOE-ε4* carrier status and tested whether and how age and healthy-lifestyle score interact with PRS to influence cognitive decline. All participants were grouped into 3 categories based on the healthy-lifestyle composite score (0-1 factors, 2-3 factors, and 4-5 factors). We also tested the association with cognitive decline with each 1-unit increase in the healthy-lifestyle score. Finally, we conducted a challenging analysis to test whether there is an interaction among *APOE*-ε4, PRS, lifestyle, and age on longitudinal cognition decline through a likelihood ratio test. All models were adjusted for sex, education, parental history of AD, practice effects (visit number minus 1), and the first five genetic principal components of ancestry. Age was centered at year 65 and education was centered at the mean of all waves for ease of interpretation. The joint significance of the three-way interaction among genetics (PRS or *APOE-ε4*), healthy-lifestyle score, and polynomial age was tested using likelihood ratio tests between the full model with and without the three-way interaction. Two-sided *P* values <0.05 after FDR correction were considered statistically significant. Once statistically significant three-way interactions were identified, we probed the nature of these interaction effects by conducting post-hoc simple-slope analyses. Specifically, we first calculated the simple-slope estimates of the genetic predictor of interest on longitudinal cognition for individuals with various numbers of healthy lifestyles from age 55 to 80 and tested them versus zero to investigate the threshold of significance. We next tested whether and when there is a statistically significant difference in the simple-slope estimates of the genetic predictor between individuals with different numbers of healthy-lifestyle factors. For all post-hoc analyses, we used an FDR-adjusted *P* value for significance to minimize false positive findings due to multiple comparisons.

We conducted a series of sensitivity analyses to evaluate the robustness of the findings. First, we adjusted for body mass index (BMI) and Center for Epidemiological Studies Depression score to estimate the impact of cardiovascular risk factors and depressive symptoms on our associations. Second, we reconstructed the healthy-lifestyle score by including only time-varying lifestyle measures (smoking, alcohol consumption, and physical activities) but with a larger sample size. Third, we leveraged the first visit at which all five lifestyle predictors were available (median visit 5) to construct time-invariant composite lifestyle measures across all waves, following the literature^23^.

Fourth, we included a healthy-lifestyle composite score by considering the relative risks of these factors from published meta-analyses (Supplementary methods)^14,17^. Fifth, we replaced the binary *APOE-ε4* carrier status with the newly introduced neuropathology *APOE* score to account for the heterogeneous effect of the ε2 and ε3 alleles in our multivariate model^30^. Sixth, we replaced the linear PRS term with linear and quadratic terms to account for the potential nonlinear effects caused by non-*APOE* genetic variants.

We also replicated our WRAP primary analyses in the Health and Retirement Study (HRS), a nationally representative longitudinal study in which the researchers have surveyed ∼42000 Americans since 1992. Since 2000, HRS has collected consistent measures on cognition^31^. Details about cognition and genetic data have been described elsewhere^31–35^. To ensure a fair comparison with WRAP participants, we only included HRS participants who were born between 1935 and 1959 (age 40 to 65 in 2000), were of genetically defined European ancestry^36^, whose cognition was not assessed through proxy, and were not classified as “demented” by the Langa-Weir Classification of Cognitive Function^34^ (Supplementary methods). Selection mortality is a known issue in the HRS genetic sample, and it may bias the gene-environment interaction results, so we separately used inverse probability weighting to account for selection mortality, as suggested by previous literature^37,38^. Due to data availability issues, we only sought replication of the findings for a healthy-lifestyle score constructed based on four factors (smoking status, physical activity, diet, and alcohol consumption; Supplementary methods).

## RESULTS

In WRAP, DNA samples were collected from 1,198 participants. Of these, 1,184 (98.8%) had non-missing cognition data in all waves, 1,163 (97.1%) had non-missing data on time-varying lifestyles (smoking, alcohol consumption, and physical activity), and 891 (74.4%) had non-missing data on visit 5 lifestyle measures on diet and cognitive activity. The final analytical sample included 891 individuals (mean baseline age of 58, 31% male, 38% *APOE-ε4* carriers; Table 1). Except for length of follow-up, there were no statistically significant differences between individuals with complete lifestyle data and the entire sample (Supplementary Table 1A). After FDR correction, we observed significant PRS×age×lifestyle interactions for delayed recall, immediate learning, and PACC-3 exclusively among *APOE-ε4* carriers. However, no significant interactions of *APOE-ε4*×age×lifestyle were observed in the full sample, and similarly, no significant *PRS*×age×lifestyle interactions were found in the full sample or among *APOE-ε4* non-carriers (Figure 1, Supplementary Tables 2A-5A). Post-hoc simple-slope analyses revealed that among *APOE-ε4* carriers, statistically significant PRS simple-slope estimates on delayed recall and immediate learning emerged after age 67 for individuals with 0-1 healthy-lifestyle factors, but no statistically significant PRS simple-slope estimates were observed among *APOE-ε4* carriers with 2-3 or 4-5 healthy-lifestyle factors across all age ranges tested (Figure 2). For PACC-3, we observed statistically significant PRS simple-slope estimates after age 67 for *APOE-ε4* carriers with 0-1 healthy-lifestyle factors and after age 72 for *APOE-ε4* carriers with 2-3 healthy-lifestyle factors. However, we observed no statistically significant PRS simple slopes among *APOE-ε4* carriers with 4-5 healthy-lifestyle factors. Next, we tested whether there was a statistically significant difference in the PRS simple-slope estimates on cognition between *APOE-ε4* carriers of the same age with different numbers of healthy-lifestyle factors at a range of ages. After FDR correction, the model parameters predicted that a significant PRS-related difference started showing after age 67-68 among *APOE-ε4* carriers with 0-1 healthy-lifestyle factors compared to more than 1 healthy-lifestyle factor on all cognitive outcomes (Figure 3). However, we observed no statistically significant difference in PRS simple slopes between *APOE-ε4* carriers with 2-3 versus 4-5 healthy-lifestyle factors across all age ranges tested. We observed the same pattern of results when we replaced the categorical adherence to healthy lifestyle measures with the continuous healthy-lifestyle composite score. Though challenging, we observed a statistically significant PRS×*APOE-ε4*×age×lifestyle on all three cognitive outcomes (all FDR-*p* < 0.01).

**Figure 1.**
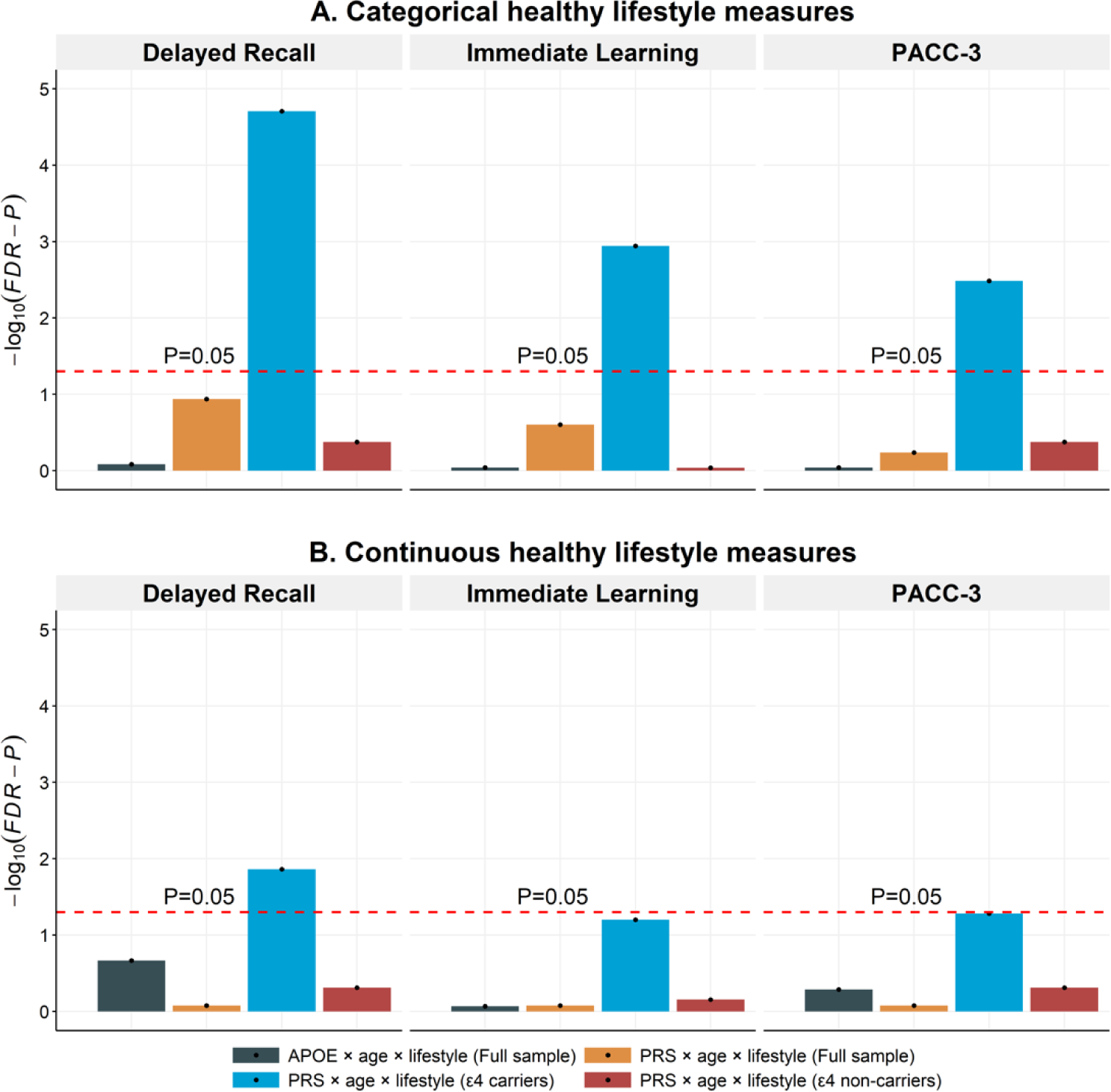
Likelihood Ratio Test (LRT) of the interactions between genetic risk predictors (*APOE-ε4* or PRS), adherence to healthy lifestyles, and age in the full sample and sample stratified by *APOE-ε4* carrier status (PRS analysis only) Figure 1 presents the FDR corrected −log_10_(*P*) from the likelihood ratio tests for the three-way interaction terms between genetic risk predictor (*APOE-ε4* or PRS), adherence to healthy lifestyles, and age in the full WRAP sample and sample stratified by *APOE-ε4* carrier status (for PRS analysis only). The likelihood ratio test statistic is calculated as the ratio between the log-likelihood of the nested model (model without three-way interaction terms) to the full model (model with genetic risk predictor*adherence to healthy lifestyles*polynomial age terms). All association tests are performed using linear mixed-effect model and adjusted for within-individual/family correlation. Additional covariates include sex, education, practice effects, family history of AD, and the first five principal components of ancestry. For the *APOE* analyses in the full sample, we additionally adjusted PRS × Age interaction in all models to control for age heterogenous genetic effect caused by non-*APOE* genetic variants. For the PRS analyses in the full sample, we additionally adjusted *APOE-ε4* × Age interaction in all models to control for age heterogenous genetic effect caused by *APOE* genetic variants, as reported previously. Age is centered at year 65 and education is centered at the mean (15.9 years). PACC-3 = Preclinical Alzheimer’s Cognitive Composite Score-3.

**Figure 2.**
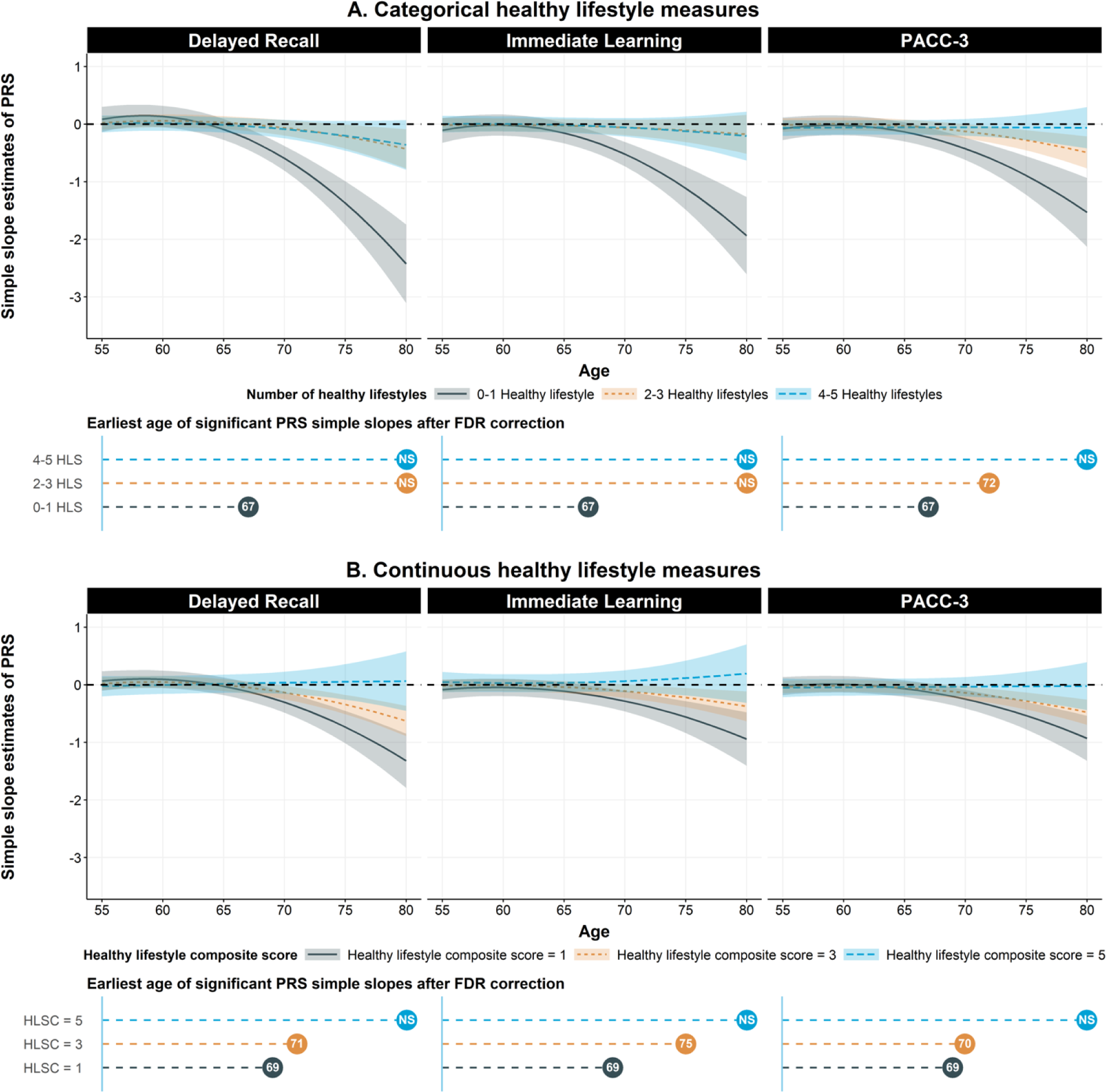
Simple slope estimates of PRS on domain specific- and global cognitive score for *APOE-ε4* carriers with different number of healthy lifestyle factors and from age 55 to 80 (N = 340). Figure 2 shows the simple slope estimates of the PRS for *APOE-ε4* carriers with different numbers of healthy lifestyles from age 55 to 80 on global and domain specific cognition score. For the categorical lifestyle measures, the grey, orange, and blue line represents the longitudinal trajectory of simple slope estimates of PRS among *APOE-ε4* carriers who have 0-1 healthy lifestyles, 2-3 healthy lifestyles, and 4-5 healthy lifestyles, respectively. For the continuous lifestyle measures (assuming linear effects of each one-unit change in healthy lifestyle), the grey, orange, and blue line represents the longitudinal trajectory of simple slope estimates of PRS among *APOE-ε4* carriers whose healthy lifestyle composite score is 1, 3, and 5, respectively. The simple slope estimates are calculated using the package “reghelper” in R and were based on the results which were obtained using the linear mixed-effect model and adjusted for within-individual/family correlation. In addition to PRS, adherence to healthy lifestyles, age (quadratic), and their interactions, additional covariates include sex, education years, practice effect, parental history of AD, and the first five principal components of ancestry. Age is centered at year 65 and education is centered at the mean (15.9 years). PACC-3 = Preclinical Alzheimer’s Cognitive Composite Score-3. HLS: Healthy lifestyles. HLSC: Healthy lifestyle composite.

**Figure 3.**
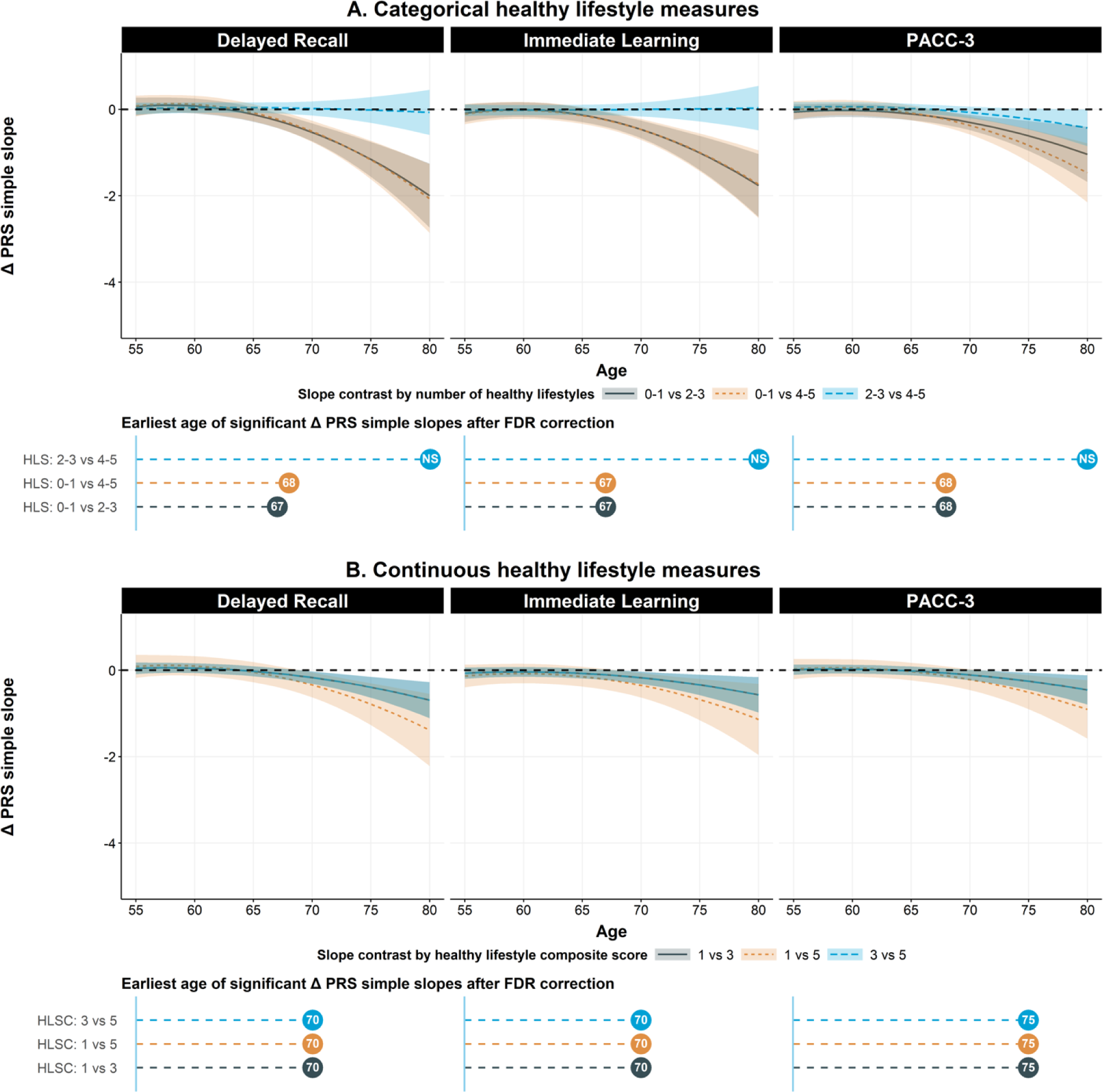
Difference in simple slope estimates of PRS on domain specific- and global cognitive score between *APOE-ε4* carriers with a different number of healthy lifestyle factors and from age 55 to 80 (N = 340). Figure 3 shows the longitudinal trajectory of the difference in simple slope estimates of PRS for *APOE-ε4* carriers who have a different number of healthy lifestyles from age 55 to 80. For categorical lifestyle measures, the grey, orange, and blue lines represent the longitudinal trajectory of the difference in simple slope estimates of PRS among *APOE-ε4* carriers: comparing those with 0-1 healthy lifestyles to those with 2-3 healthy lifestyles, those with 0-1 healthy lifestyles to those with 4-5 healthy lifestyles, and those with 2-3 healthy lifestyles to those with 4-5 healthy lifestyles, respectively. Concerning continuous lifestyle measures (assuming linear effects of each one-unit change in healthy lifestyle), the grey, orange, and blue lines represent the longitudinal trajectory of the difference in simple slope estimates of PRS among *APOE-ε4* carriers: comparing individuals with a healthy lifestyle composite score of 1 to those with a score of 3, individuals with a score of 1 to those with a score of 5, and individuals with a score of 3 to those with a score of 5, respectively. The difference in simple slope estimates were calculated using the package “emmeans” in R and were based on the results that were obtained using the linear mixed-effect model and adjusted for within-individual/family correlation. In addition to PRS, age (quadratic), adherence to healthy lifestyles, and their interactions, additional covariates include sex, education years, practice effect, parental history of AD, and the first five principal components of ancestry. Age is centered at year 65 and education is centered at the mean (15.9 years). PACC-3 = Preclinical Alzheimer’s Cognitive Composite Score-3. HLS: Healthy lifestyles. HLSC: Healthy lifestyle composite.

**Table 1.** Descriptive statistics of WRAP sample at visit 2 data collection.

| Variables | N = 891<sup>1</sup> |
| --- | --- |
| Age | 58 (6) |
| Maximum visits<sup>2</sup> | 5.03 (0.78) |
| Education | 15.91 (2.24) |
| Family history of AD | 663 / 891 (74%) |
| Male | 277 / 891 (31%) |
| Current smoker | 44 / 891 (4.9%) |
| **Drinking habit** |  |
| Current non-drinker | 150 / 891 (17%) |
| Current light drinker | 261 / 891 (29%) |
| Current moderate drinker | 273 / 891 (31%) |
| Current heavy drinker | 207 / 891 (23%) |
| **Exercise habit** |  |
| < 150 min/week | 510 / 891 (57%) |
| 150-300 min/week | 210 / 891 (24%) |
| 300+ min/week | 171 / 891 (19%) |
| MIND<sup>3</sup> diet score at visit 5 | 8.64 (1.84) |
| FCAS<sup>4</sup> Cognitive activity score at visit 5 | 44 (8) |
| **APOE genotype** |  |
| ε2/ε2 | 2 / 891 (0.2%) |
| ε2/ε3 | 75 / 891 (8.4%) |
| ε3/ε3 | 474 / 891 (53%) |
| ε2/ε4 | 30 / 891 (3.4%) |
| ε3/ε4 | 276 / 891 (31%) |
| ε4/ε4 | 34 / 891 (3.8%) |
| CESD<sup>5</sup> score | 6 (6) |
| BMI | 28.7 (5.8) |

<sup>1</sup>Mean (SD); n / N (%)
<sup>2</sup> "Maximum visits" refers to number of cognitive assessments an individual contributed to these analyses.
<sup>3</sup> MIND: Mediterranean-DASH Diet Intervention for Neurodegenerative Delay
<sup>4</sup> FCAS: Florida Cognitive Activities Scale
<sup>5</sup> CESD: Center for Epidemiologic Studies Depression Scale

In sensitivity analyses, we observed similar patterns to those found in the main analysis when we further adjusted for BMI and CESD score as covariates (Supplementary Figure 1), used the time-invariant lifestyle measure constructed based on the first visit at which all five lifestyle predictors were available (Supplementary Figure 2), and reconstructed and categorized the healthy-lifestyle score by considering these factors’ relative risks (Supplementary Figure 3). We also observed similar findings when we replaced the *APOE-ε4* carriers’ status with the continuous *APOE* score and used nonlinear PRS terms (results not shown). However, models using the healthy-lifestyle measure constructed based on the three time-varying healthy-lifestyle factors showed an attenuated modification effect compared to the main analyses (Supplementary Figure 4).

In the HRS replication analyses, we observed similar patterns to those from the main analyses for delayed recall and global cognition, whether considering all available healthy lifestyles (N=1,864, Supplementary Figures 5-7) or only time-varying healthy lifestyles (N=6,686, Supplementary Figures 8-9). However, HRS participants with diet data available are generally healthier than the general population (Supplementary Table 1B). Furthermore, we noted significant interactions among PRS, age, and adherence to a healthy lifestyle in the full sample and among *APOE-ε4* noncarriers although the modifying effects were considerably weaker compared to *APOE-ε4* carriers. Significant PRS×*APOE*-ε4×age×lifestyle interaction was also observed on delayed recall and global cognition. Nonetheless, models that do not account for mortality selection generally displayed attenuated modification effects. Similar results were also observed in the sensitivity analyses in the HRS dataset (Supplementary methods, results not shown).

## DISCUSSION

Utilizing two longitudinal studies, WRAP and HRS, we discovered that healthy-lifestyle factors moderate the risk of known non-*APOE* genetic variants on longitudinal cognitive decline, with protective effects more pronounced among *APOE-ε4* carriers. To our knowledge, no prior study has investigated this four-way interaction effect.

Past epidemiological studies have investigated how lifestyle choices affect the genetic risk of LOAD, yielding mixed results ^15,16,39,40^, with some null findings even in a large sample^15^. Recent research has demonstrated that age modifies LOAD genetic risk ^18,41,42^ and that PRS effects on LOAD-related traits are stronger among *APOE-ε4* carriers^22,43–45^. However, these findings have not been integrated into previous gene×lifestyle interaction analyses, as the present analysis does.

In this study, we also considered lifestyle over time in constructing the composite lifestyle measures. In HRS and WRAP, our results confirm that a favorable lifestyle can mitigate the adverse genetic effects caused by non-*APOE* common genetic variants, particularly among *APOE-ε4* carriers. Although we did observe several significant interactions between lifestyle, age, and PRS/*APOE-ε4* in the full sample analyses, the overall protective effects of adhering to healthy lifestyles are notably more pronounced for PRS among *APOE-ε4* carriers. Also, we observed a potential threshold for the protective effects of adherence to healthy lifestyles. In both HRS and WRAP, the protective effects became apparent with more than one healthy lifestyles.

Two findings warrant further investigation with a larger sample size, extended follow-up, and a more comprehensive lifestyle database. First, adhering to a healthy lifestyle might influence when genetic effects become noticeable throughout one’s life. Prior work suggested adverse PRS effects emerged by age 70 in *APOE-ε4* carriers^22^. The present analysis in the same cohort suggests that among *APOE-ε4* carriers with the least healthy habits, cognitive decline begins approximately 1-3 years earlier, although this was not replicated in HRS. Second, in the WRAP dataset, statistically significant interactions were observed among PRS, lifestyle, and age, but only in *APOE-ε4* carriers. In contrast, in the HRS dataset, these interactions were significant in the full sample, albeit more pronounced among *APOE-ε4* carriers. As previously reported, a subtle trend of PRS effects was detected among *APOE-ε4* noncarriers in the HRS dataset but not in WRAP^19^.

### Limitations

Our study has several limitations. First, its power is limited by the availability of healthy-lifestyle data, even in HRS, especially when we stratify the sample by *APOE-ε4* carrier status. Nevertheless, we were able to demonstrate the robustness of our findings in several sensitivity analyses in both cohorts. Second, of the five lifestyle factors we included, two are time invariant and do not capture lifestyle changes during follow-up. Third, lifestyle factors are self-reported and are subject to recall and social desirability bias. Fourth, in the current study, we only considered the combined influence of five healthy-lifestyle factors based on established evidence; however, other lifestyle factors may also be involved in the etiology of LOAD. Fifth, our results could be subject to bias caused by unmeasured confounding and reverse causation due to the long prodromal phase of LOAD. Sixth, our study is limited to individuals of European ancestry and may not be generalizable to other populations. Seventh, diet data in HRS were collected from 2013 to 2014, and HRS participants with available diet data are generally healthier than the general population, affecting generalizability and potentially biasing estimates toward the null. Eighth, without precise evaluation of the cause of cognitive decline, we couldn’t confirm if the observed interaction effects are truly part of LOAD etiology. This limitation might make it difficult to replicate our results in other community-based samples, especially when participants are more likely affected by specific health conditions related to cognitive decline, such as vascular disease. Ninth, the protective effects of living healthily against the genetic risk of LOAD might be due to brain resilience, reduced inflammation, or other biological pathways. However, these questions cannot be addressed without replicating the analysis in a well-powered biomarker sample. Tenth, at baseline, the consensus conference process for assigning cognitive status had not started, and there might have been a handful of mild cognitive impairment cases at baseline that were included in the study.

## CONCLUSION

Among individuals without cognitive impairment or dementia, the effect of LOAD genetic risk on longitudinal cognitive decline may depend on lifestyle factors. These protective modifying effects are particularly pronounced among *APOE-ε4* carriers.

## Data Availability

All data produced in the present study are available upon reasonable request to the authors.

## Author Contributions

Dr Engelman and Mr. Xu had full access to all of the data in the study and takes responsibility for the integrity of the data and the accuracy of the data analysis.

*Concept and design:* Engelman, Xu.

*Acquisition, analysis, or interpretation of data*: Engelman, Xu, Johnson.

*Drafting of the manuscript:* Engelman, Xu.

*Critical review of the manuscript for important intellectual content*: Engelman, Xu, Sun, Jonaitis, Deming, Lu, Johnson.

*Statistical analysis*: Xu, Sun.

*Obtained funding:* Engelman.

*Administrative, technical, or material support:* Engelman, Johnson.

*Supervision:* Engelman.

## Conflict of Interest Disclosures

Dr. Johnson has served as a consultant to Roche Diagnostics, Merck and Prothena and has received research funding from Cerveau Technologies. No other disclosures were reported.

## Funding/Support

This study was supported by the National Institutes of Health (NIH) grants R01AG27161 (Wisconsin Registry for Alzheimer Prevention: Biomarkers of Preclinical AD), R01AG054047 (Genomic and Metabolomic Data Integration in a Longitudinal Cohort at Risk for Alzheimer’s Disease), and R21AG067092 (Identifying Metabolomic Risk Factors in Plasma and Cerebrospinal Fluid for Alzheimer’s Disease), the Helen Bader Foundation, Northwestern Mutual Foundation, Extendicare Foundation, State of Wisconsin, the Clinical and Translational Science Award (CTSA) program through the NIH National Center for Advancing Translational Sciences (NCATS) grant [UL1TR000427]. Computational resources were supported by core grants to the Center for Demography and Ecology [P2CHD047873] and the Center for Demography of Health and Aging [P30AG017266]. Author YX was supported by a Center for Demography of Health and Aging pilot award [P30AG017266]. The HRS (Health and Retirement Study) is sponsored by the National Institute on Aging (grant number NIA U01AG009740) and is conducted by the University of Michigan. The HRS genetic data were collected with financial support from the National Institute of Health’s (NIH) Director’s Opportunity for Research awards using American Reinvestment and Recovery Act funds (RC2 AG036495-01, RC4 AG039029-01) and were accessed through NIAGADS with accession number NG00119.v1.

## Role of the Funder/Sponsor

The funders had no role in the design and conduct of the study; collection, management, analysis, and interpretation of the data; preparation, review, or approval of the manuscript; and decision to submit the manuscript for publication.

## Data Sharing Statement

See Supplement 2.

## Notes

### Author Declarations

The University of Wisconsin-Madison Institutional Review Board gave ethical approval for this work.

