## Supplementary material for "Mid-to-Late Life Healthy Lifestyle Modifies Genetic Risk for Longitudinal Cognitive Aging among Asymptomatic Individuals": Supplmentary methods and figures

[1]G.H. Sergievsky Center, Vagelos College of Physicians and Surgeons, Columbia University; [2]Department of Biostatistics and Medical Informatics, School of Medicine and Public Health, University of Wisconsin-Madison; [3]Department of Medicine, School of Medicine and Public Health, University of Wisconsin-Madison; [4]Wisconsin Alzheimer's Institute, University of Wisconsin-Madison; [5]Wisconsin Alzheimer's Disease Research Center, University of Wisconsin-Madison

*Correspondence:*
Corinne D. Engelman, MSPH, PhD
Department of Population Health Sciences,
University of Wisconsin - Madison
610 Walnut Street, 1007A WARF
Madison, WI 53726-2397


Yuexuan Xu, MPP, PhD
The Gertrude H. Sergievsky Center
Columbia University
630 W 168th Street, PH19 303
New York, NY 10032


Yuexuan Xu will handle correspondence at all stages of refereeing and publication, also post-publication.

### SUPPLEMENTARY MATERIALS

#### *Table of Contents for Supplementary Methods and Figures*

**Supplementary Figure 5. Likelihood Ratio Test (LRT) of the interactions between genetic risk predictors (*APOE* ε4 or PRS), adherence to healthy lifestyles (smoking,**

#### *Table of Contents for Supplementary Tables*

- Supplementary Table 1A. Descriptive statistics of WRAP sample at visit 2 data collection, and comparisons of the observed characteristics between WRAP participants with three-lifestyle factors (full sample) and five lifestyle factors (survive from visit 2 until at least visit 5)

- Supplementary Table 1B. Descriptive statistics of HRS sample at first visit, and comparisons of the observed characteristics between HRS participants with three-lifestyle factors (full sample) and four lifestyle factors (survive from 2000 until at least 2014)

- Supplementary Table 2A. Associations between APOE ε4, adherence to healthy lifestyle, and age on global and domain-specific cognition score in WRAP sample (Five lifestyles, N = 891)

- Supplementary Table 2B. Associations between APOE ε4, adherence to healthy lifestyle, and age on global and domain-specific cognition score in HRS sample (Four lifestyles, N = 1,864)

- Supplementary Table 3A. Associations between PRS, adherence to healthy lifestyle, and age on global and domain-specific cognition score in WRAP sample (Five lifestyles, N = 891)

- Supplementary Table 3B. Associations between PRS, adherence to healthy lifestyle, and age on global and domain-specific cognition score in HRS sample (Four lifestyles, N = 1,864)

- Supplementary Table 4A. Associations between PRS, adherence to healthy lifestyle, and age on global and domain-specific cognition score in among APOE ε4 carriers in WRAP (Five lifestyles, N = 340)

- Supplementary Table 4B. Associations between PRS, adherence to healthy lifestyle, and age on global and domain-specific cognition score in among APOE ε4 carriers in HRS (Five lifestyles, N = 447)

- Supplementary Table 5A. Associations between PRS, adherence to healthy lifestyle, and age on global and domain-specific cognition score in among APOE ε4 non-carriers in WRAP (Five lifestyles, N = 551)

- Supplementary Table 5B. Associations between PRS, adherence to healthy lifestyle, and age on global and domain-specific cognition score in among APOE ε4 non-carriers in HRS (Five lifestyles, N = 1,417)

### SUPPLEMENTARY METHODS IN WRAP

#### Assessment of healthy lifestyles and other covariates in WRAP

Lifestyle measures included in the current study encompass smoking status, physical activity, diet, alcohol consumption, and cognitive activity. Information on smoking status was obtained through questionnaire interviews at each study visit, where participants specified whether they had smoked any cigarettes at all in the past month. If yes, then this individual was identified as a current smoker. Physical activity was measured using the Women's Health Initiative physical activities questionnaire in WRAP[1]. We calculated the weekly time (in minutes) that WRAP participants engaged in moderate or vigorous physical activities based on their responses to questions about the frequency and duration of moderate (e.g., calisthenics) and vigorous (e.g., jogging) physical activities.

Alcohol intake was calculated as the amount per week based on respondents' answers to a questionnaire about the number of days per week they consumed alcoholic beverages and how many drinks they typically consumed per day on the days they drank. We then categorized WRAP participants into current non-drinker, current light drinker, current moderate drinker, and current heavy drinker based on the guidelines provided by the National Health Interview Survey (NHIS)[2].

Dietary intake was assessed by the Mediterranean-DASH Intervention for Neurodegenerative Delay Questionnaire (MIND; 15 items)[3]. The MIND diet score is based on the consumption of 10 "brain-healthy" food groups (e.g., leafy greens, nuts, berries, fish) and 5 unhealthy food groups (e.g., red meats, fried food, pastries/sweets). Since we evaluated alcohol intake separately, we did not include the MIND measure for the average frequency of intake of alcoholic beverages into the MIND diet score calculation, following the literature[4].

Cognitive activity was measured by a total cognitive activities score, which is constructed based on respondents' answers to the Florida Cognitive Activities Scale (FCAS), including 25 items covering a range of cognitive activities (e.g., playing games, solving puzzles)[5].

Depressive symptoms were assessed with the Center for Epidemiologic Studies Depression (CESD) scale.

#### Classification of healthy lifestyle categories

Following previous literature[4], a healthy lifestyle was classified as follows: (1) not currently smoking, (2) engaging in moderate or vigorous exercise activities for at least 150 minutes per week, (3) consuming alcohol in light to moderate amounts, (4) having a MIND diet score (without alcohol) in the top 40% of the cohort distribution, and (5) achieving a total cognitive activities score in the top 40% of the cohort distribution.

| Lifestyle factor | Low-risk category | High-risk category |
|---|---|---|
| Alcohol consumption | Light or moderate drinker | Non-drinker and heavy drinkers |
| Physical exercises | ≥150 min/week in moderate or vigorous activities | <150 min/week or sedentary |
| Smoking habits | Current non-smoker | Current smoker |
| Cognitive activity | Upper 2/5ths (highest 40%) of the distribution | Bottom 3/5ths (lower 60%) of the distribution |

| | | |
|---|---|---|
| MIND Diet score | Upper 2/5ths (highest 40%) of the distribution | Bottom 3/5ths (lower 60%) of the distribution |

#### Incorporating relative risk of healthy lifestyles into the calculation of healthy lifestyle score

We followed the method for constructing the Lifestyle for Brain Health (LIBRA) index and created another weighted healthy lifestyle score by additionally incorporating the relative risk of each individual healthy lifestyle factor[6,7]. Specifically, we calculated this weighted sum score by assigning weights to each factor, which were obtained from the published meta-analysis and are in accordance with the relative risks. In the sensitivity analysis, we categorized the weighted sum score into three groups based on tertiles and also used it as a continuous variable for the analysis, following the literature[7].

| Healthy lifestyle | Operationalization | Weight |
|---|---|---|
| Current non-smoking | Self-reported current smokers or non-smokers. | +1.5 |
| Low-to-moderate alcohol use | Self-reported weekly frequency of alcohol consumed | +1.0 |
| Physical active | Self-reported moderate/vigorous activities > 150 min/week. | +1.1 |
| High cognitive activity | FCAS cognitive activities total score | +3.2 |
| Healthy diet | MIND Diet score | +1.7 |

#### Simple effects of *APOE* ε4 calculation

We first calculated the predicted value of cognition for individuals based on individuals' *APOE* ε4 carrier status and number of healthy lifestyles from age 55 to 80. At each single age, there are six combinations between *APOE* ε4 carrier status and number of healthy (use categorical lifestyle as an example):

- *APOE* ε4 carrier with 0-1 healthy lifestyle
- *APOE* ε4 carrier with 2-3 healthy lifestyle
- *APOE* ε4 carrier with 4-5 healthy lifestyle
- *APOE* ε4 non-carrier with 0-1 healthy lifestyle
- *APOE* ε4 non-carrier with 2-3 healthy lifestyle
- *APOE* ε4 non-carrier with 4-5 healthy lifestyle

To compute the simple effect of *APOE* ε4, we take the difference in the predicted value of cognition for individuals who are *APOE* ε4 carrier and non-carriers but have the same number of healthy lifestyles and at the same age. The difference in simple effects of *APOE* ε4 is calculated as the difference in the difference in the predicted value of cognition for individuals who are *APOE* ε4 carrier and non-carriers but have the same number of healthy lifestyles and at the same age.

#### Simple slopes of PRS calculation (CATEGORICAL lifestyle measure)

Suppose we have a three-way interaction model with categorical lifestyle measures (0-1, 2-3, and 4-5), using individuals with 0-1 healthy lifestyle as the reference.

$Y = \beta_0 + \beta_1 PRS + \beta_2 Age + \beta_3 Age^2 + \beta_4 Lifestyle(2\text{-}3) + \beta_5 Lifestyle(4\text{-}5) + \beta_6 Age*PRS + \beta_7 Age^2*PRS + \beta_8 Age*Lifestyle(2\text{-}3) + \beta_9 Age*Lifestyle(4\text{-}5) + \beta_{10} Age^2*Lifestyle(2\text{-}3) +$

$\beta_{11}Age^2*Lifestyle(4\text{-}5) + \beta_{12}PRS*Lifestyle(2\text{-}3) + \beta_{13}PRS*Lifestyle(4\text{-}5) + \beta_{14}PRS*Lifestyle(2\text{-}3)*Age + \beta_{15}PRS*Lifestyle(4\text{-}5)*Age + \beta_{16}PRS*Lifestyle(2\text{-}3)*Age^2 + \beta_{17}PRS*Lifestyle(4\text{-}5)*Age^2 + BX + i + f + u$

Where *BX* represents covariates (gender, education, parental history of AD, practice effects, and first five principal components), i index the within-individual random intercept, j is the within-family random intercept, and u is the error term.

To calculate the simple slope of the *PRS* for individuals with different *number of healthy lifestyles* at different age, we can reorganize the above model as

$Y = \beta_0 + PRS(\beta_1 + \beta_6Age + \beta_7Age^2 + \beta_{12}Lifestyle(2\text{-}3) + \beta_{13}Lifestyle(4\text{-}5) + \beta_{14}Lifestyle(2\text{-}3)*Age + \beta_{15}Lifestyle(4\text{-}5)*Age + \beta_{16}Lifestyle(2\text{-}3)*Age^2 + \beta_{17}Lifestyle(4\text{-}5)*Age^2) +$
$\beta_2Age + \beta_3Age^2 + \beta_4Lifestyle(2\text{-}3) + \beta_5Lifestyle(4\text{-}5) + \beta_8Age*Lifestyle(2\text{-}3) + \beta_9Age*Lifestyle(4\text{-}5) + \beta_{10}Age^2*Lifestyle(2\text{-}3) + \beta_{11}Age^2*Lifestyle(4\text{-}5) + BX + i + f + u$

Then it's easy to calculate the simple slope of PRS by using different combination of values for age and *APOE* ε4 in this three-way interaction model.

#### EXAMPLE

If we want to calculate the simple slope of PRS on PACC-3 for *APOE ε4 carriers* with different number of healthy lifestyles and at age 65, 70, and 75.

We first get the regression outputs as the following (the same as the model presented in table 1, age is centered at year 65)

| | PACC-3 | | |
|---|---|---|---|
| *Predictors* | *Estimates* | *std. Error* | *p* |
| (β0) (Intercept) | -0.96123909 | 0.18115034 | <0.001 |
| (β1) PRS | -0.1353355 | 0.08339584 | 0.105 |
| (β2) Age (centered at year 65) | -0.07164843 | 0.01379105 | <0.001 |
| (β3) $Age^2$ (centered at year 65) | -0.001913 | 0.00099589 | 0.055 |
| (β4) 2-3 Healthy lifestyles | 0.04395247 | 0.08260623 | 0.595 |
| (β5) 4-5 Healthy lifestyles | 0.0788586 | 0.09441329 | 0.404 |
| (β6) PRS * Age | -0.04062438 | 0.00967906 | <0.001 |
| (β7) PRS * $Age^2$ | -0.00350112 | 0.00092385 | <0.001 |
| (β8) 2-3 Healthy lifestyles * Age | -0.01052323 | 0.01223961 | 0.39 |
| (β9) 4-5 Healthy lifestyles * Age | -0.00403818 | 0.01315685 | 0.759 |
| (β10) 2-3 Healthy lifestyles * $Age^2$ | -0.00037825 | 0.00103914 | 0.716 |
| (β11) 4-5 Healthy lifestyles * $Age^2$ | 0.00005164 | 0.00109231 | 0.962 |
| (β12) 2-3 Healthy lifestyles * PRS | 0.10466284 | 0.07345409 | 0.154 |
| (β13) 4-5 Healthy lifestyles * PRS | 0.0799398 | 0.08661171 | 0.356 |
| (β14) 2-3 Healthy lifestyles * PRS * Age | 0.02758379 | 0.01029566 | 0.007 |
| (β15) 4-5 Healthy lifestyles * PRS * Age | 0.04096662 | 0.01128763 | <0.001 |

| | | | |
|---|---|---|---|
| (β16) 2-3 Healthy lifestyles * PRS * Age$^2$ | 0.00232503 | 0.00100382 | 0.021 |
| (β17) 4-5 Healthy lifestyles * PRS * Age$^2$ | 0.00344009 | 0.00105264 | 0.001 |
| Family history of AD | 0.01908856 | 0.14587354 | 0.896 |
| Female | 0.64534616 | 0.1027308 | <0.001 |
| Education | 0.1321006 | 0.02247455 | <0.001 |
| Practice effects | 0.08039707 | 0.02097562 | <0.001 |
| PC1 | -2.27722899 | 1.93156289 | 0.239 |
| PC2 | 1.88886964 | 1.77537396 | 0.288 |
| PC3 | 2.83133766 | 1.67068203 | 0.09 |
| PC4 | -0.02679995 | 1.66700726 | 0.987 |
| PC5 | 2.61353156 | 1.75609165 | 0.137 |

The simple slopes of PRS on PACC-3 for individuals with 0-1 healthy lifestyles and at age 65, 70, 75 (0, 5, 10 for age-65-centered age) are

Age 65:

$-0.1353355 + \beta_6*0 + \beta_7*0 + \beta_{12}*0 + \beta_{13}*0 + \beta_{14}*0 + \beta_{15}*0 + \beta_{16}*0 + \beta_{17}*0 =$
$-0.1353355$

Age 70:

$-0.1353355 + \beta_6*5 + \beta_7*25 + \beta_{12}*0 + \beta_{13}*0 + \beta_{14}*0 + \beta_{15}*0 + \beta_{16}*0 + \beta_{17}*0 =$
$-0.1353355 + (-0.04062438)*5 + (-0.00350112)*25 = -0.4259854$

Age 75:

$-0.1353355 + \beta_6*10 + \beta_7*100 + \beta_{12}*0 + \beta_{13}*0 + \beta_{14}*0 + \beta_{15}*0 + \beta_{16}*0 + \beta_{17}*0 =$
$-0.1353355 + (-0.04062438)*10 + (-0.00350112)*100 = -0.8916913$

The simple slopes of PRS on PACC-3 for individuals with 2-3 healthy lifestyles and at age 65, 70, 75 (0, 5, 10 for age-65-centered age) are

Age 65:

$-0.1353355 + \beta_6*0 + \beta_7*0 + \beta_{12}*1 + \beta_{13}*0 + \beta_{14}*0 + \beta_{15}*0 + \beta_{16}*0 + \beta_{17}*0 =$
$-0.1353355 + 0.10466284 = -0.03067266$

Age 70:

$-0.1353355 + \beta_6*5 + \beta_7*25 + \beta_{12}*1 + \beta_{13}*0 + \beta_{14}*1*5 + \beta_{15}*0 + \beta_{16}*1*25 + \beta_{17}*0 =$
$-0.1353355 + (-0.04062438)*5 + (-0.00350112)*25 + 0.10466284 + 0.02758379*5 + 0.00232503*25 = -0.1252779$

Age 75:

$-0.1353355 + \beta_6*10 + \beta_7*100 + \beta_{12}*1 + \beta_{13}*0 + \beta_{14}*1*10 + \beta_{15}*0 + \beta_{16}*100 + \beta_{17}*0 =$

-0.1353355 + (-0.04062438)*10 + (-0.00350112)*100 + 0.10466284 + 0.02758379*10 + 0.00232503*100 = -0.2786876

The simple slopes of PRS on PACC-3 for individuals with 4-5 healthy lifestyles and at age 65, 70, 75 (0, 5, 10 for age-65-centered age) are

<u>Age 65:</u>

-0.1353355 + $\beta_6*0 + \beta_7*0 + \beta_{12}*0 + \beta_{13}*1 + \beta_{14}*0 + \beta_{15}*0 + \beta_{16}*0 + \beta_{17}*0$ =
-0.1353355 + 0.0799398 = -0.0553957
<u>Age 70</u>

-0.1353355 + $\beta_6*5 + \beta_7*25 + \beta_{12}*0 + \beta_{13}*1 + \beta_{14}*0 + \beta_{15}*5 + \beta_{16}*0 + \beta_{17}*25$ =
-0.1353355 + (-0.04062438)*5 + (-0.00350112)*25 + 0.0799398 + 0.04096662*5 + 0.00344009*25 = -0.05521025

<u>Age 75:</u>

-0.1353355 + $\beta_6*10 + \beta_7*100 + \beta_{12}*0 + \beta_{13}*1 + \beta_{14}*0 + \beta_{15}*10 + \beta_{16}*0 + \beta_{17}*100$ =
-0.1353355 + (-0.04062438)*10 + (-0.00350112)*100 + 0.0799398 + 0.04096662*10 + 0.00344009*100 = -0.0580763

#### Simple slopes of PRS calculation (CONTINUOUS lifestyle measure)

Suppose we have a three-way interaction model with continuous healthy lifestyle measures (healthy lifestyle composite: range 0-5),

$Y = \beta_0 + \beta_1 PRS + \beta_2 Age + \beta_3 Age^2 + \beta_4 Lifestyle + \beta_5 Age*PRS + \beta_6 Age^2*PRS + \beta_7 Age*Lifestyle + \beta_8 Age^2*Lifestyle + \beta_9 PRS*Lifestyle + \beta_{10} PRS*Lifestyle*Age + \beta_{11} PRS*Lifestyle*Age^2 + BX + i + f + u$
Where *BX* represents covariates (gender, education, parental history of AD, practice effects, and first five principal components), i index the within-individual random intercept, j is the within-family random intercept, and u is the error term.

To calculate the simple slope of the *PRS* for individuals with different *APOE* ε4 carrier status at different age, we can reorganize the above model as

$Y = \beta_0 + PRS(\beta_1 + \beta_5 Age + \beta_6 Age^2 + \beta_9 Lifestyle + \beta_{10} Lifestyle*Age + \beta_{11} Lifestyle*Age^2) + \beta_2 Age + \beta_3 Age^2 + \beta_4 APOE + \beta_7 Age*APOE + \beta_8 Age^2*APOE + BX + i + f + u$

Then it's easy to calculate the simple slope of PRS by using different combination of values for age and healthy lifestyle score in this three-way interaction model.

EXAMPLE

If we want to calculate the simple slope of PRS on PACC-3 for <u>*APOE ε4 carriers*</u> with different number of healthy lifestyles and at age 65, 70, and 75.

We first get the regression outputs as the following (the same as the model presented in table 1, age is centered at year 65)

| | PACC-3 | | |
| --- | --- | --- | --- |
| *Predictors* | *Estimates* | *std. Error* | *p* |
| (β0) (Intercept) | -1.027528159 | 0.184201913 | <0.001 |
| (β1) PRS | -0.073525287 | 0.091779028 | 0.423 |
| (β2) Age (centered at year 65) | -0.096477539 | 0.011869272 | <0.001 |
| (β3) Age$^2$(centered at year 65) | -0.003105646 | 0.000782199 | <0.001 |
| (β4) Healthy lifestyle score | 0.038324249 | 0.026482168 | 0.148 |
| (β5) PRS * Age | -0.03241301 | 0.008948092 | <0.001 |
| (β6) PRS * Age$^2$ | -0.002647747 | 0.000818141 | 0.001 |
| (β7) Healthy lifestyle score * Age | 0.005237889 | 0.002987838 | 0.08 |
| (β8) Healthy lifestyle score * Age$^2$ | 0.000290325 | 0.000245863 | 0.238 |
| (β9) Healthy lifestyle score * PRS | 0.007187322 | 0.025786649 | 0.78 |
| (β10) Healthy lifestyle score * PRS * Age | 0.006719265 | 0.002820083 | 0.017 |
| (β11) Healthy lifestyle score * PRS * Age$^2$ | 0.000526746 | 0.000261132 | 0.044 |
| Family history of AD | 0.022729688 | 0.145770947 | 0.876 |
| Female | 0.643416446 | 0.102669748 | <0.001 |
| Education | 0.130552957 | 0.022458835 | <0.001 |
| Practice effects | 0.080022395 | 0.020966589 | <0.001 |
| PC1 | -2.219350648 | 1.93035718 | 0.25 |
| PC2 | 1.978893771 | 1.774805749 | 0.265 |
| PC3 | 2.871956842 | 1.671216999 | 0.086 |
| PC4 | 0.003474123 | 1.665883481 | 0.998 |
| PC5 | 2.674413217 | 1.754938594 | 0.128 |

The simple slopes of PRS on PACC-3 for individuals with individuals whose healthy lifestyle composite score = 1 and at age 65, 70, 75 (0, 5, 10 for age-65-centered age) are

<u>Age 65:</u>

-0.073525287+ $\beta_5*0 + \beta_6*0 + \beta_9*1 + \beta_{10}*0 + \beta_{11}*0 =$
-0.073525287+ 0.007187322 = -0.06633796

<u>Age 70:</u>

-0.073525287+ $\beta_5*5 + \beta_6*25 + \beta_9*1 + \beta_{10}*1*5 + \beta_{11}*1*25 =$
-0.073525287+ (-0.03241301)*5 + (-0.002647747)*25 + 0.007187322 + 0.006719265*1*5 + 0.000526746*1*25 = -0.2478317

<u>Age 75:</u>

-0.073525287+ $\beta_5*10 + \beta_6*100 + \beta_9*1 + \beta_{10}*1*10 + \beta_{11}*1*100 =$
-0.073525287+ (-0.03241301)*10 + (-0.002647747)*100 + 0.007187322 + 0.006719265*1*10 + 0.000526746*1*100 = -0.5353755

The simple slopes of PRS on PACC-3 for individuals with individuals whose healthy lifestyle composite score = 3 and at age 65, 70, 75 (0, 5, 10 for age-65-centered age) are

Age 65:

-0.073525287+ $\beta_5*0 + \beta_6*0 + \beta_9*3 + \beta_{10}*0 + \beta_{11}*0$ =
-0.073525287+ 0.007187322*3 = -0.05196332

Age 70:

-0.073525287+ $\beta_5*5 + \beta_6*25 + \beta_9*3 + \beta_{10}*3*5 + \beta_{11}*3*25$ =
-0.073525287+ (-0.03241301)*5 + (-0.002647747)*25 + 0.007187322*3 + 0.006719265*3*5 + 0.000526746*3*25 = -0.1399271

Age 75:

-0.073525287+ $\beta_5*10 + \beta_6*100 + \beta_9*3 + \beta_{10}*3*10 + \beta_{11}*3*100$ =
-0.073525287+ (-0.03241301)*10 + (-0.002647747)*100 + 0.007187322*3 + 0.006719265*3*10 + 0.000526746*3*100 = -0.2812664

The simple slopes of PRS on PACC-3 for individuals with individuals whose healthy lifestyle composite score = 5 and at age 65, 70, 75 (0, 5, 10 for age-65-centered age) are

Age 65:

-0.073525287+ $\beta_5*0 + \beta_6*0 + \beta_9*5 + \beta_{10}*0 + \beta_{11}*0$ =
-0.073525287+ 0.007187322*5 = -0.03758868

Age 70:

-0.073525287+ $\beta_5*5 + \beta_6*25 + \beta_9*5 + \beta_{10}*5*5 + \beta_{11}*5*25$ =
-0.073525287+ (-0.03241301)*5 + (-0.002647747)*25 + 0.007187322*5 + 0.006719265*5*5 + 0.000526746*5*25 = -0.03202253

Age 75:

-0.073525287+ $\beta_5*10 + \beta_6*100 + \beta_9*5 + \beta_{10}*5*10 + \beta_{11}*5*100$ =
-0.073525287+ (-0.03241301)*10 + (-0.002647747)*100 + 0.007187322*5 + 0.006719265*5*10 + 0.000526746*5*100 = -0.02715723

#### Likelihood ratio test for the joint significance of the interaction effects

We test the joint significance of the interactions between PRS, adherence to healthy lifestyle (healthy lifestyle composite score), and age using likelihood ratio tests. Specifically, we tested the coefficients for all the three-way interaction terms are simultaneously zero (degree of freedom = difference between number of parameters in the full model vs nested model).

**Categorical healthy lifestyle measure:**

Full model:

$$Y= \beta_0 + \beta_1 PRS + \beta_2 Age + \beta_3 Age^2 + \beta_4 Lifestyle(2\text{-}3) + \beta_5 Lifestyle(4\text{-}5) + \beta_6 Age*PRS + \beta_7 Age^2*PRS + \beta_8 Age*Lifestyle(2\text{-}3) + \beta_9 Age*Lifestyle(4\text{-}5) + \beta_{10} Age^2*Lifestyle(2\text{-}3) + \beta_{11} Age^2*Lifestyle(4\text{-}5) + \beta_{12} PRS*Lifestyle(2\text{-}3) + \beta_{13} PRS*Lifestyle(4\text{-}5) + \beta_{14} PRS*Lifestyle(2\text{-}3)*Age + \beta_{15} PRS*Lifestyle(4\text{-}5)*Age + \beta_{16} PRS*Lifestyle(2\text{-}3)*Age^2 + \beta_{17} PRS*Lifestyle(4\text{-}5)*Age^2 + BX + i + f + u \text{ (30 terms)}$$

Nested model:

$$Y= \beta_0 + \beta_1 PRS + \beta_2 Age + \beta_3 Age^2 + \beta_4 Lifestyle(2\text{-}3) + \beta_5 Lifestyle(4\text{-}5) + \beta_6 Age*PRS + \beta_7 Age^2*PRS + \beta_8 Age*Lifestyle(2\text{-}3) + \beta_9 Age*Lifestyle(4\text{-}5) + \beta_{10} Age^2*Lifestyle(2\text{-}3) + \beta_{11} Age^2*Lifestyle(4\text{-}5) + \beta_{12} PRS*Lifestyle(2\text{-}3) + \beta_{13} PRS*Lifestyle(4\text{-}5) + BX + i + f + u \text{ (26 terms)}$$

Where *BX* represents covariates (gender, education, family history of AD, practice effects, and first five principal components), i index the within-individual random intercept, j is the within-family random intercept, and u is the error term.  Age*PRS/Age**APOE* was adjusted only in the PRS or *APOE* analysis in the full sample.

**Continuous healthy lifestyle measure:**

Full model:

$$Y= \beta_0 + \beta_1 PRS + \beta_2 Age + \beta_3 Age^2 + \beta_4 Lifestyle + \beta_5 Age*PRS + \beta_6 Age^2*PRS + \beta_7 Age*Lifestyle + \beta_8 Age^2*Lifestyle + \beta_9 PRS*Lifestyle + \beta_{10} PRS*Lifestyle*Age + \beta_{11} PRS*Lifestyle*Age^2 + BX + i + f + u \text{ (24 terms)}$$

Nested model:

$$Y= \beta_0 + \beta_1 PRS + \beta_2 Age + \beta_3 Age^2 + \beta_4 Lifestyle + \beta_5 PRS*Lifestyle + \beta_6 Lifestyle*Age + \beta_7 Lifestyle*Age^2 + \beta_8 PRS*Age + \beta_9 PRS*Age^2 + BX + i + f + u \text{ (22 terms)}$$

Where *BX* represents covariates (gender, education, family history of AD, practice effects, and first five principal components), i index the within-individual random intercept, j is the within-family random intercept, and u is the error term. Age*PRS/Age**APOE* was adjusted only in the PRS or *APOE* analysis in the full sample.

#### Better understand simple slopes via predicted values plot

Another way to visualize simple slopes is to plot predicted values based on individuals' different numbers of lifestyles, age, and different levels of genetic risk. In the current study, we didn't create predicted plots because of the large number of figures that would be produced. For example, if we wanted to plot a three-way interaction between age, lifestyle, and PRS among *APOE* ε4 carriers, using five-year age intervals from 55 to 80, we would need to create at least six figures for a single cognitive outcome for each model. To save space in this manuscript, we therefore present the longitudinal trajectory of simple slopes/effects because it can more efficiently convey the results.

We provide an example below to illustrate the connection between simple slope estimates in the predicted value plot. We used PACC-3 as the outcome and plotted the predicted cognitive values for individuals with healthy lifestyle composite scores of 1, 3, and 5, respectively, using five-year age intervals.

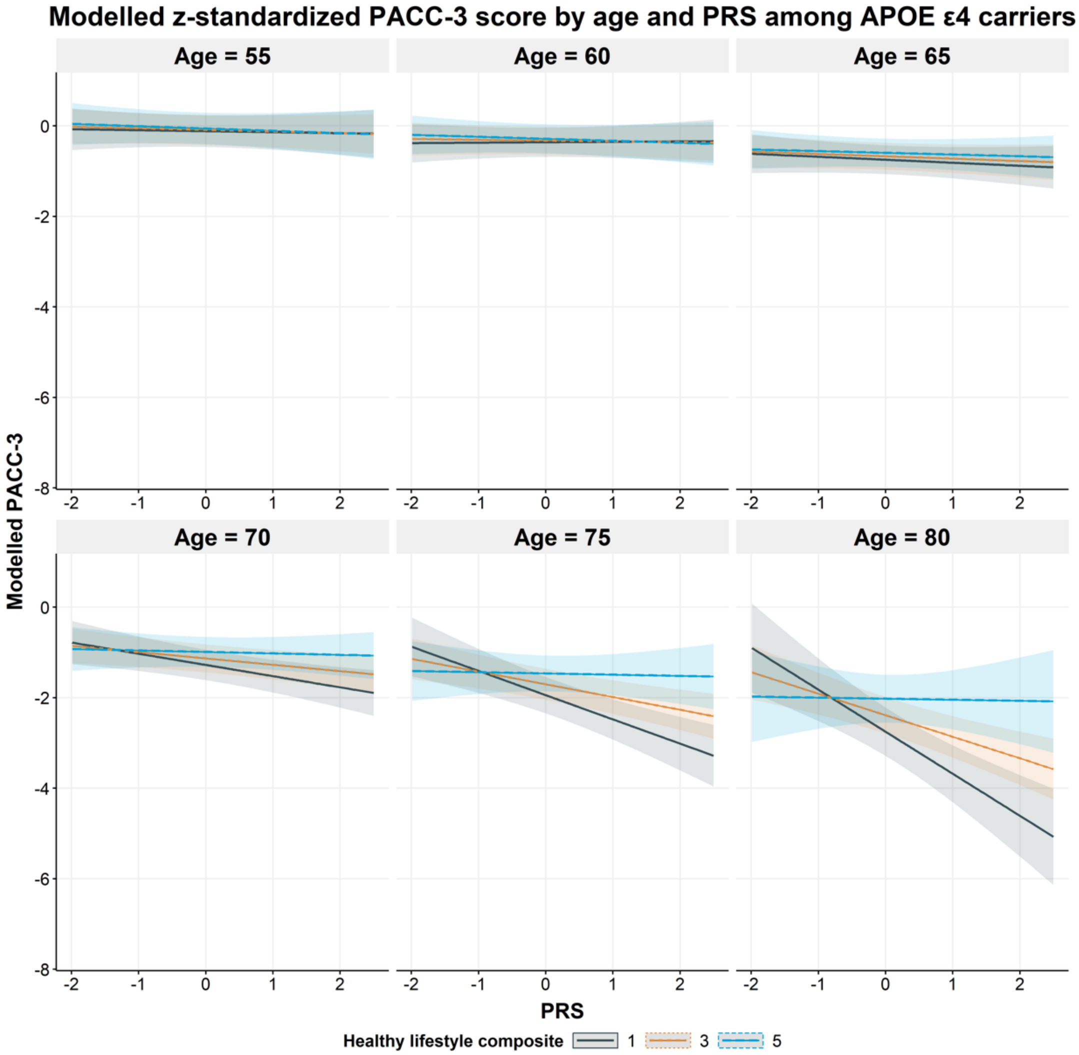

Corresponding to the simple slope estimates of PRS we have manually calculated above among *APOE* ε4 carriers, the slope of the grey line for individuals whose healthy lifestyle composite score is 1 at age 65, 70, and 75 is -0.066338, -0.247831, and -0.535376, respectively.

The slope of the orange line for individuals whose healthy lifestyle composite score is 3 at age 65, 70, and 75 is -0.0519633, -0.139927, and -0.281266, respectively.

The slope of the blue line for individuals whose healthy lifestyle composite score is 3 at age 65, 70, and 75 is -0.0375889, -0.0320226, and -0.0271556, respectively.

### SUPPLEMENTARY METHODS IN HRS

#### Assessment of healthy lifestyles and other covariates in HRS

Lifestyle measures included in the HRS replication analysis were extracted from the RAND HRS longitudinal file 2018 (v2) (N ≈ 42,000) and the HRS Health Care and Nutrition (HCNS) sub-study conducted between November 2013 and May 2014[8]. Of the five healthy lifestyles analyzed in the WRAP original study, four were also available in the HRS, including smoking status, physical activity, diet, and alcohol consumption. Diet data were collected cross-sectionally and are only available in a subset of HRS participants (N = 8,035).

Smoking status information was collected through questionnaire interviews at each study visit. Participants indicated whether they were current smokers or non-smokers. Being a current non-smoker was considered a protective lifestyle factor, while being a current smoker was considered a risk lifestyle factor.

Due to the absence of data regarding the duration of weekly physical exercises, we were unable to create a measure representing the weekly time HRS participants spent on moderate or vigorous physical activities. Instead, we adhered to guidelines from previous literature, focusing solely on the weekly frequency of participating in moderate or vigorous physical activities. Consequently, we defined regular physical activity as engaging in moderate physical activity on at least two days a week or vigorous activity at least once a week[9].

We calculated alcohol intake based on respondents' answers to a questionnaire that inquired about the number of days per week they consumed alcoholic beverages and the quantity consumed per day on drinking days. We categorized HRS participants into four groups—current non-drinker, current light drinker, current moderate drinker, and current heavy drinker—using the guidelines from the National Health Interview Survey (NHIS)[2].

Dietary intake was assessed using the Mediterranean-DASH Intervention for Neurodegenerative Delay Questionnaire (MIND; 15 items)[3,8]. The MIND diet score is based on the consumption of 10 'brain-healthy' food groups (e.g., leafy greens, nuts, berries, fish) and 5 unhealthy food groups (e.g., red meats, fried food, pastries/sweets). We excluded alcohol intake from the MIND diet score calculation, as it was evaluated separately, in line with existing literature. Diet data are available only for a subsample of HRS participants[8].

Depressive symptoms were assessed with the Center for Epidemiologic Studies Depression (CESD) scale.

#### Classification of healthy lifestyle categories

| Lifestyle factor | Low-risk category | High-risk category |
|---|---|---|
| Alcohol consumption | Light or moderate drinker | Non-drinker and heavy drinkers |
| Physical exercises | ≥2 days/week in moderate or >= 1 days/week vigorous activities | < 2 days/week in moderate and < 1 days/week vigorous activities |
| Smoking habits | Current non-smoker | Current smoker |
| Cognitive activity | NA | NA |
| MIND Diet score | Upper 2/5ths (highest 40%) of the distribution | Bottom 3/5ths (lower 60%) of the distribution |

#### Replication analysis in HRS

We were unable to fully replicate our WRAP findings in HRS due to significant differences in data collection methods for lifestyle measures between the two studies. For instance, HRS only collects information on the frequency of participating in moderate/vigorous exercises per week and does not collect data on the duration of weekly exercises. Additionally, some lifestyle data are only available in a subsample of HRS participants. For example, dietary data are only accessible for a subset of HRS participants (8,035 out of 42,000, approximately 1/5 of the total sample), and including diet data in the composite measure would result in a reduction of the sample size to 1/5 of its original size after applying exclusion criteria. As a result, we have developed various composite measures for healthy lifestyles by incorporating as much information as possible while simultaneously assessing the sensitivity of our findings to different combinations of healthy lifestyles, which include –

- Smoking, alcohol consumption, physical exercises, and diet (main replication analyses)
- Smoking, alcohol consumption, physical exercises (test for the sensitivity of the exclusion of diet data, main replication analyses)
- Smoking, alcohol consumption, diet (test for the sensitivity of the exclusion of physical exercises data, results not shown, available upon request)
- Smoking, alcohol consumption (test for the sensitivity of the exclusion of both diet and physical exercises data, results not shown, available upon request)

#### Genotyping, *APOE,* and non-*APOE* PRS in HRS

Genotype data on over 15,000 HRS participants were obtained using the Illumina HumanOmni2.5 BeadChip[10]. Individuals with missing call rates >2% or chromosomal anomalies were excluded. Also, SNPs that do not meet the quality control criteria, including intensity-only or duplicate SNPs, SNPs with MAF = 0, missing call rate ≥2%, >2 discordant calls in 103 study duplicates, >1 Mendelian error, Hardy-Weinberg equilibrium p-value <1e-4, sex difference in allelic frequency ≥0.2, and sex difference in heterozygosity >0.3 were excluded. Genotype data were imputed to a worldwide 1000 Genomes Project reference panel using SHAPEIT2 [11] and IMPUTE2 [12] software. Genotype imputation was performed and documented by the Genetics Coordinating Center at the University of Washington. Only SNPs that were directly genotyped or imputed with info score >0.8 were kept in the analysis. We replicated the WRAP main analyses by using PRS constructed based on the genome-wide significant variants as identified by Kunkle et al[13] as the main polygenic predictor and used the same procedure of constructing PRS as that described in the WRAP analyses. *APOE* genotype was first divided into six sub-genotypes (ε2/ε2, ε2/ε3, ε3/ε3, ε2/ε4, ε3/ε4, and ε4/ε4) based on rs7412 and rs429358 and then combined into two groups that include individuals who are *APOE* ε4 carriers (ε2/ε4, ε3/ε4, ε4/ε4) and non-carriers (ε2/ε2, ε2/ε3, ε3/ε3).

#### Covariates and outcomes in HRS

Since 2000, HRS has collected consistent measures on cognition with a 27-point modified version of the Telephone Interview for Cognitive Status (TICSm)[14–17]. The TICSm assesses immediate recall (0-10 points) and delayed recall (0-10 points) tests for memory performance, serial 7s subtraction (0-5 points) tests of working memory, and backwards counting from 20 (0-2 points), which was designed to measure processing speed. Details about these tests have been described elsewhere[18]. A global cognition composite score was created by summing all the items within the tests mentioned above, with a maximum of 27 points. In the replication analysis, we used data from the 2000 wave of the HRS with follow-up until 2018. To ensure a fair comparison to the WRAP results, we focused on the preclinical stage of AD by only including HRS participants who were born between 1935 and 1959 (age 40 to 65 at year 2000), were genetically determined as European descent[19], whose cognition was not assessed through proxy, and were not classified as "demented" by the Langa-Weir Classification of Cognitive Function[15].

All other statistical methods in the replication analyses were the same as those described in the WRAP analysis, except for the exclusion of parental history of AD as a covariate because measures on family history of AD were collected after 2010[20]. We also included an indicator for cohort as a covariate because HRS enrolled a new cohort every six years and this measure was adjusted by the other AD-related HRS longitudinal cognition analyses[21]. To make the HRS replication analyses comparable to the WRAP findings, we additionally standardized all the cognitive outcomes with a mean of 0 and standard deviation of 1.

#### Accounting for selection mortality in HRS genetic sample

Selection mortality is a known issue in the HRS genetic sample because HRS genetic data were collected starting in 2006, while other demographic, health, and retirement data were collected beginning in 1992. It is recommended that formal techniques, such as inverse probability weighting, be employed to correct for selection when analyzing the interactions between genetic risk factors and environments in HRS[22,23].

To construct inverse probability weights for mortality selection, we first estimated the probability of survival (being included) into our genetic sample using a logistic regression:

$$\Pr(Person\ i\ dies\ 2006\ or\ larer\ |\ X_i) = \frac{\exp(X_i'\beta)}{1+\exp(X_i'\beta)}$$

For the predictor matrix *X*, we considered a set of health indicators which include years of education, mean BMI across all waves, maximum height across all waves, ever smoker, ever have diabetes, ever have heart disease, mean CESD score across all waves, mean self-reported health of the respondent across all waves, gender, and birth year, following previous literature[22].

Results from the logistic regressions were used to estimate the weights for the inverse probability weighting. We used inverse probability weighting to weight the observed sample to be more reflective of the sample prior to mortality selection.

### SUPPLEMENTARY FIGURES

**Supplementary Figure 1. Simple slope estimates of PRS on domain specific- and global cognitive score for *APOE* ε4 carriers with different number of healthy lifestyles and from age 55 to 80, after the adjustment of additional covariates (N = 340).**

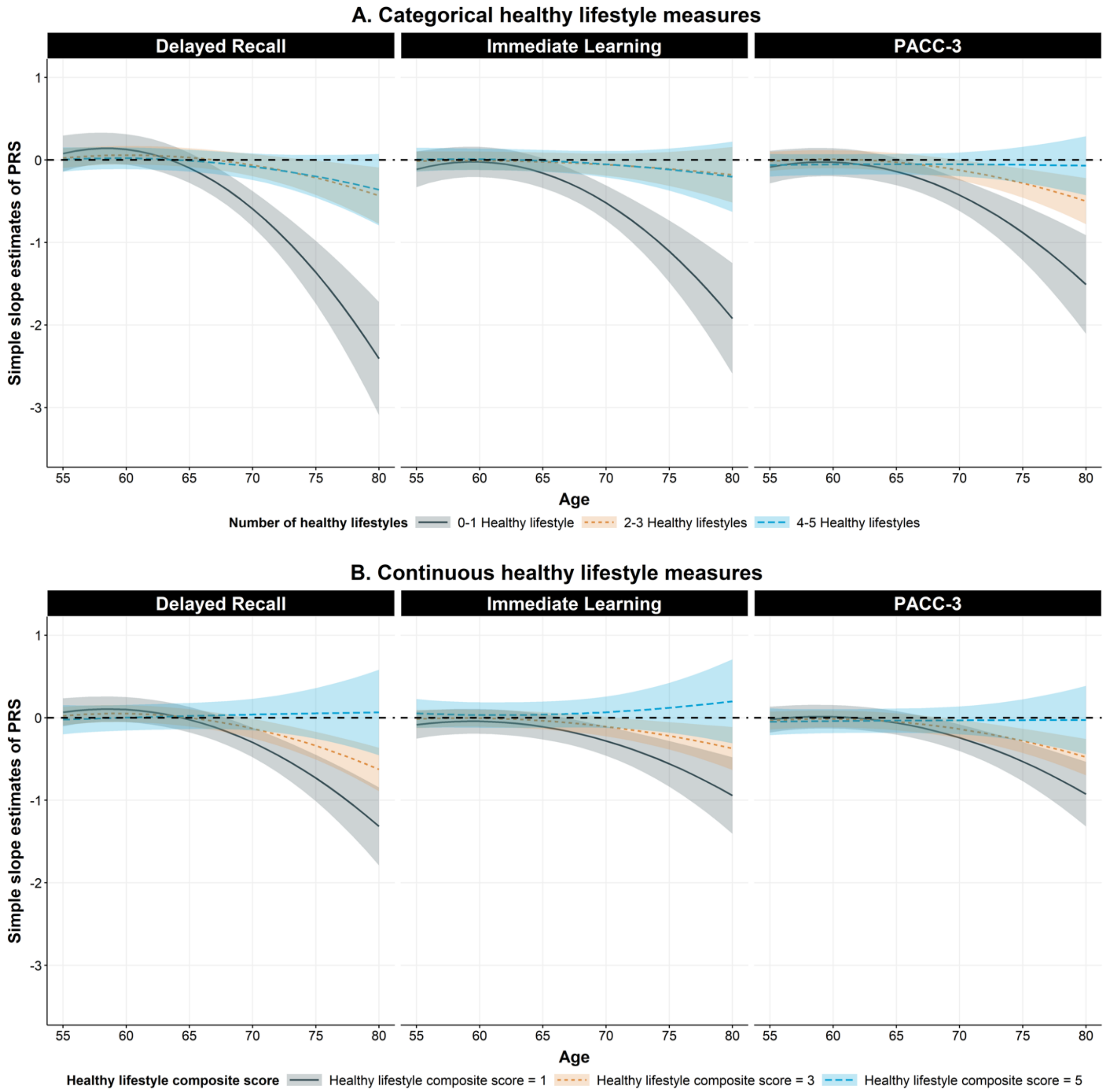

Supplementary Figure 1 shows the simple slope estimates of the PRS for *APOE* ε4 carriers with a different number of healthy lifestyles from age 55 to 80 on global and domain specific cognition score, after the adjustment of additional cardiovascular risk factors (e.g., BMI) and comorbid conditions (e.g., depression). For the categorical lifestyle measures, the grey, orange, and blue line represents the longitudinal trajectory of simple slope estimates of PRS among *APOE* ε4 carriers who have 0-1 healthy lifestyles, 2-3 healthy lifestyles, and 4-5 healthy lifestyles, respectively. For the continuous lifestyle measures, the grey, orange, and blue line represents the longitudinal trajectory of simple slope estimates of PRS among *APOE* ε4 carriers whose healthy lifestyle composite score is 1, 3, and 5, respectively. The simple slope estimates are calculated using the package "reghelper" in R and were based on the results which were obtained using the linear mixed-effect model and adjusted for within-individual/family correlation. In addition to PRS, age (quadratic), adherence to healthy lifestyles, and their interactions, additional covariates include sex, education years, practice effect, parental history of AD, BMI, CESD-score, and the first five principal components of ancestry. Age is centered at year 65 and education is centered at the mean (15.9 years). PACC-3 = Preclinical Alzheimer's Cognitive Composite Score-3. CESD-score = Center for Epidemiologic Studies Depression Scale score.

**Supplementary Figure 2. Simple slope estimates of PRS on domain specific- and global cognitive score for *APOE* ε4 carriers with different number of healthy lifestyles and from age 55 to 80, when the lifestyle measures was constructed based on the data at the first visit at which all healthy lifestyle factors were available (median visit = 5) (N = 340).**

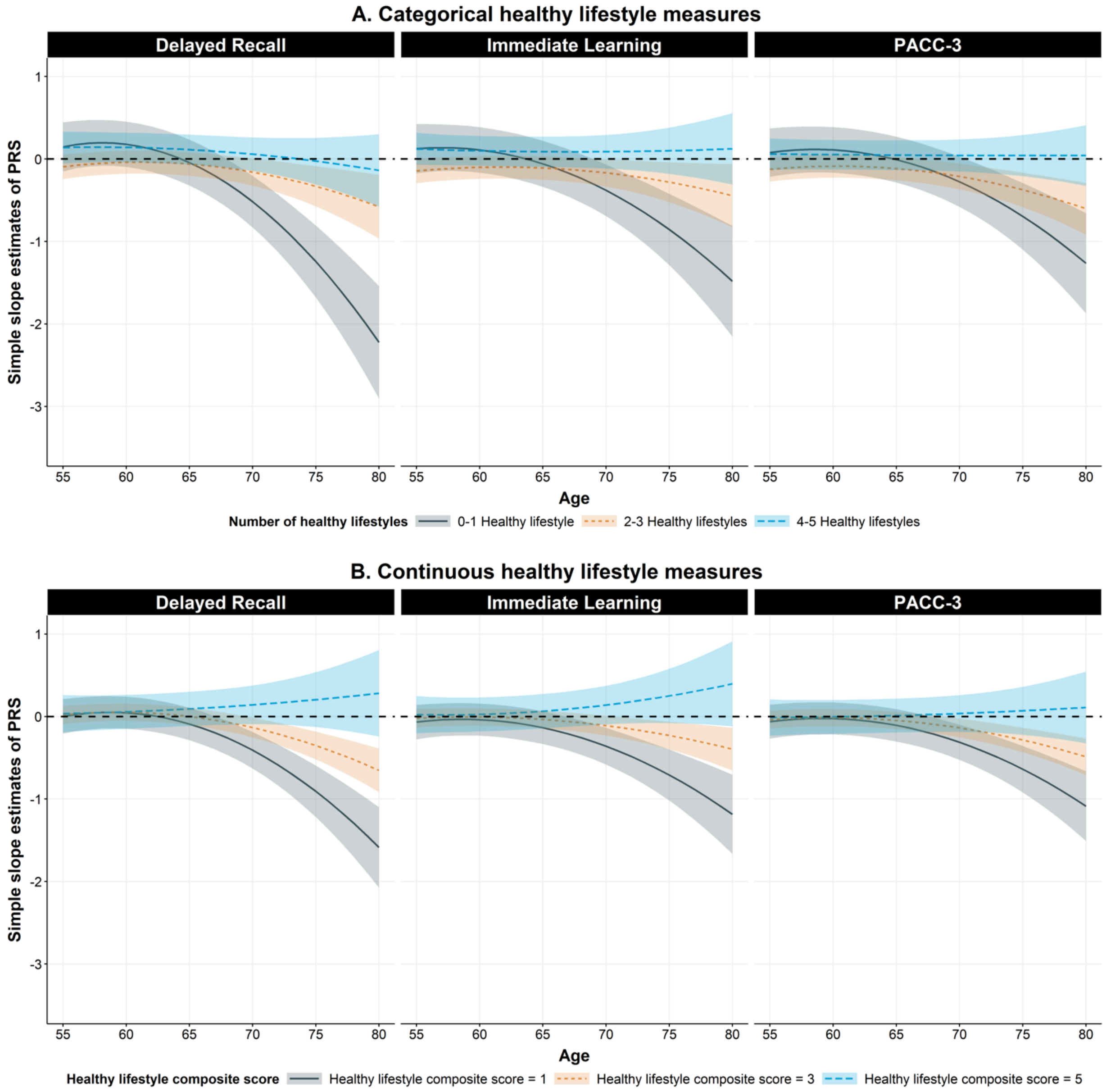

Supplementary Figure 2 shows the simple slope estimates of the PRS for *APOE* ε4 carriers with a different number of healthy lifestyles from age 55 to 80 on global and domain specific cognition score. The measure for the combined influence of healthy lifestyles was constructed using data from the first visit where all healthy lifestyle factors were available, with a median visit of 5. For the categorical lifestyle measures, the grey, orange, and blue line represents the longitudinal trajectory of simple slope estimates of PRS among *APOE* ε4 carriers who have 0-1 healthy lifestyles, 2-3 healthy lifestyles, and 4-5 healthy lifestyles, respectively. For the continuous lifestyle measures, the grey, orange, and blue line represents the longitudinal trajectory of simple slope estimates of PRS among *APOE* ε4 carriers whose healthy lifestyle composite score is 1, 3, and 5, respectively. The simple slope estimates were calculated using the package “reghelper” in R and were based on the results which were obtained using the linear mixed-effect model and adjusted for within-individual/family correlation. In addition to PRS, age (quadratic), adherence to healthy lifestyles, and their interactions, additional covariates include sex, education years, practice effect, parental history of AD, and the first five principal components of ancestry. Age is centered at year 65 and education is centered at the mean (15.9 years). PACC-3 = Preclinical Alzheimer’s Cognitive Composite Score-3.

**Supplementary Figure 3. Simple slope estimates of PRS on domain specific- and global cognitive score for *APOE* ε4 carriers with different number of healthy lifestyles and from age 55 to 80, after considering the relative risk of each lifestyle factor (N = 340).**

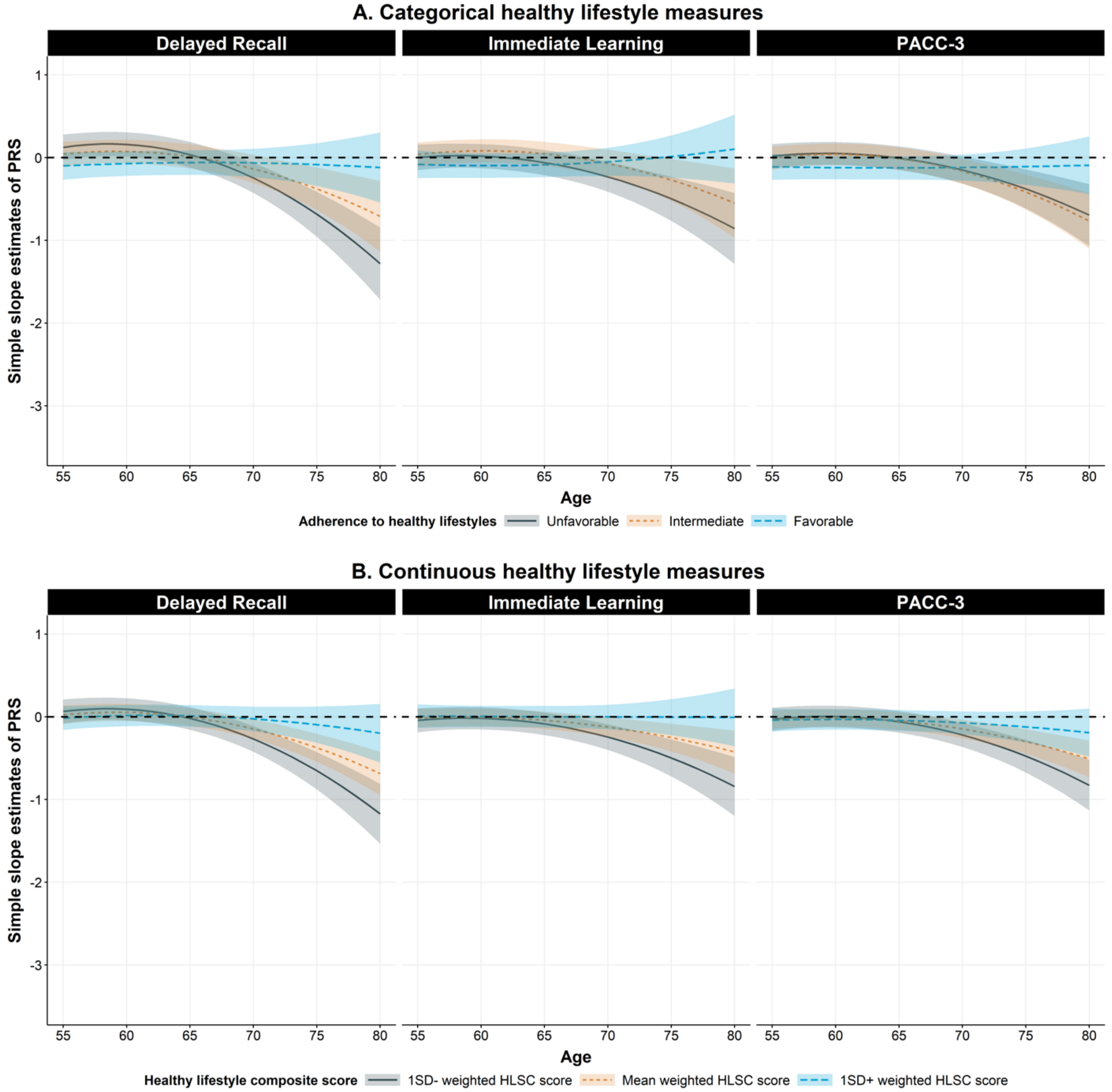

Supplementary Figure 3 shows the simple slope estimates of the PRS for *APOE* ε4 carriers with a different number of healthy lifestyles from age 55 to 80 on global and domain specific cognition score, after considering the relative risk of each lifestyle factor. We calculated this weighted sum score by assigning weights to each factor, which were obtained from the published meta-analysis and are in accordance with the relative risks. A higher weighted score represents a healthier lifestyle. We then categorized the weighted sum score into three groups based on tertiles and also used it as a continuous variable for the analysis. For the categorical lifestyle measures, the grey, orange, and blue line represents the longitudinal trajectory of simple slope estimates of PRS for individuals who have unfavorable (lowest tertile of the score distribution), intermediate (middle tertile of the score distribution), and favorable (upper tertile of the score distribution) lifestyle, respectively. For the continuous lifestyle measures, the grey, orange, and blue line represents the longitudinal trajectory of simple slope estimates of PRS for individuals whose healthy lifestyle composite score is 1 SD-, at the mean, and 1 SD+ in the score distribution, respectively. The simple slope estimates are calculated using the package "reghelper" in R and were based on the results which were obtained using the linear mixed-effect model and adjusted for within-individual/family correlation. In addition to PRS, age (quadratic), adherence to healthy lifestyles, and their interactions, additional covariates include sex, education years, practice effect, parental history of AD, and the first five principal components of ancestry. Age is centered at year 65 and education is centered at the mean (15.9 years). PACC-3 = Preclinical Alzheimer's Cognitive Composite Score-3.

**Supplementary Figure 4. Simple slope estimates of PRS on domain specific- and global cognitive score for *APOE* ε4 carriers with a different number of healthy lifestyles and from age 55 to 80, when only considering alcohol consumption, smoking, and physical exercises (N = 451).**

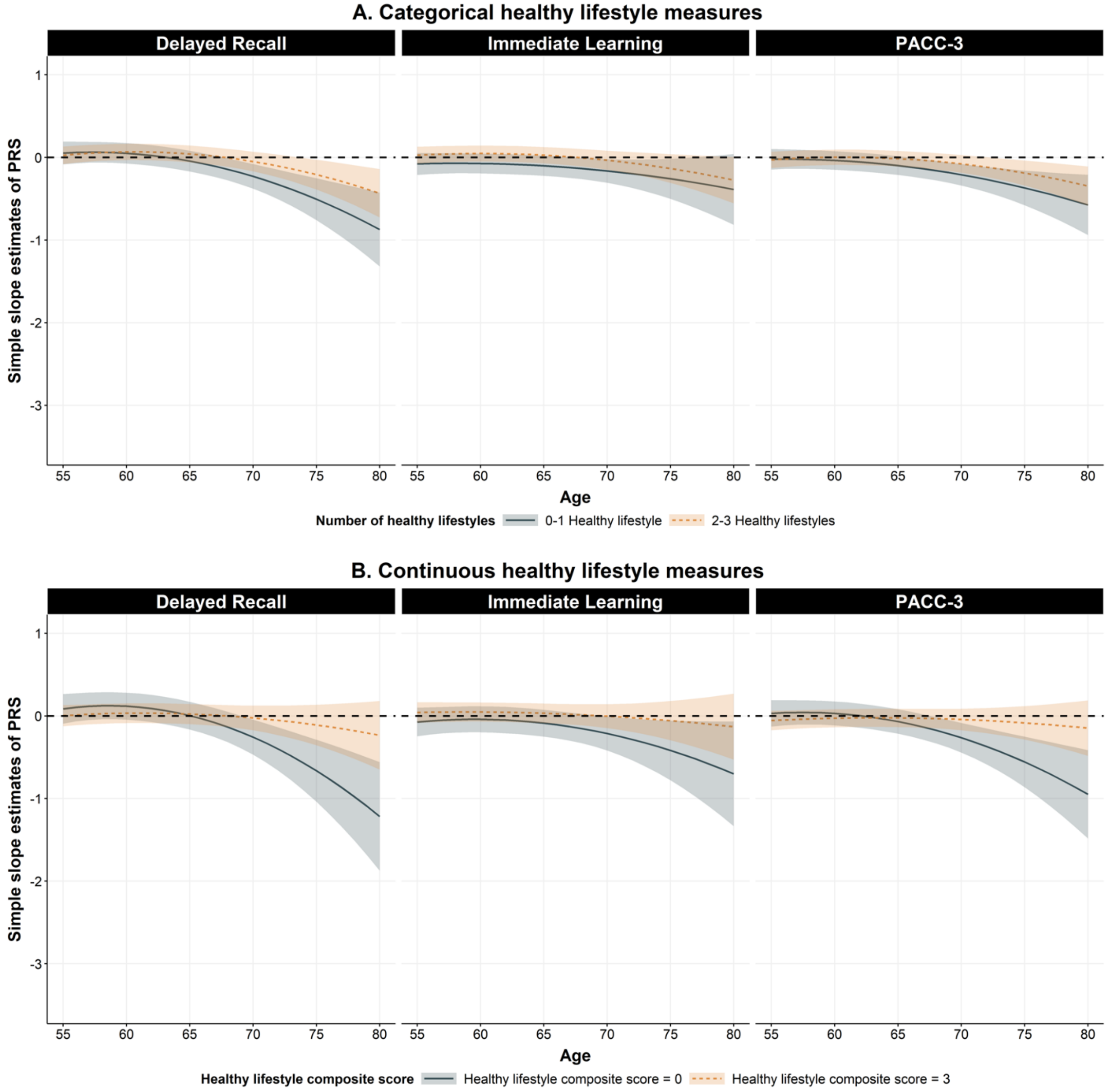

Supplementary Figure 4 shows the simple slope estimates of the PRS for *APOE* ε4 carriers with a different number of healthy lifestyles from age 55 to 80 on global and domain specific cognition score. For the categorical lifestyle measures, the grey and orange line represents the longitudinal trajectory of simple slope estimates of PRS among *APOE* ε4 carriers who have 0-1 healthy lifestyles and 2-3 healthy lifestyles, respectively. For the continuous lifestyle measures, the grey and orange line represents the longitudinal trajectory of simple slope estimates of PRS among *APOE* ε4 carriers whose healthy lifestyle composite score is 0 and 3, respectively. The simple slope estimates are calculated using the package “reghelper” in R and were based on the results which were obtained using the linear mixed-effect model and adjusted for within-individual/family correlation. In addition to PRS, age (quadratic), adherence to healthy lifestyles, and their interactions, additional covariates include sex, education years, practice effect, parental history of AD, and the first five principal components of ancestry. Age is centered at year 65 and education is centered at the mean (15.8 years). PACC-3 = Preclinical Alzheimer's Cognitive Composite Score-3.

**Supplementary Figure 5. Likelihood Ratio Test (LRT) of the interactions between genetic risk predictors (*APOE* ε4 or PRS), adherence to healthy lifestyles (smoking, physical activity, diet, and alcohol consumption), and Age in the full sample and sample stratified by *APOE* ε4 carrier status in HRS (PRS analysis only) (N = 1,864)**

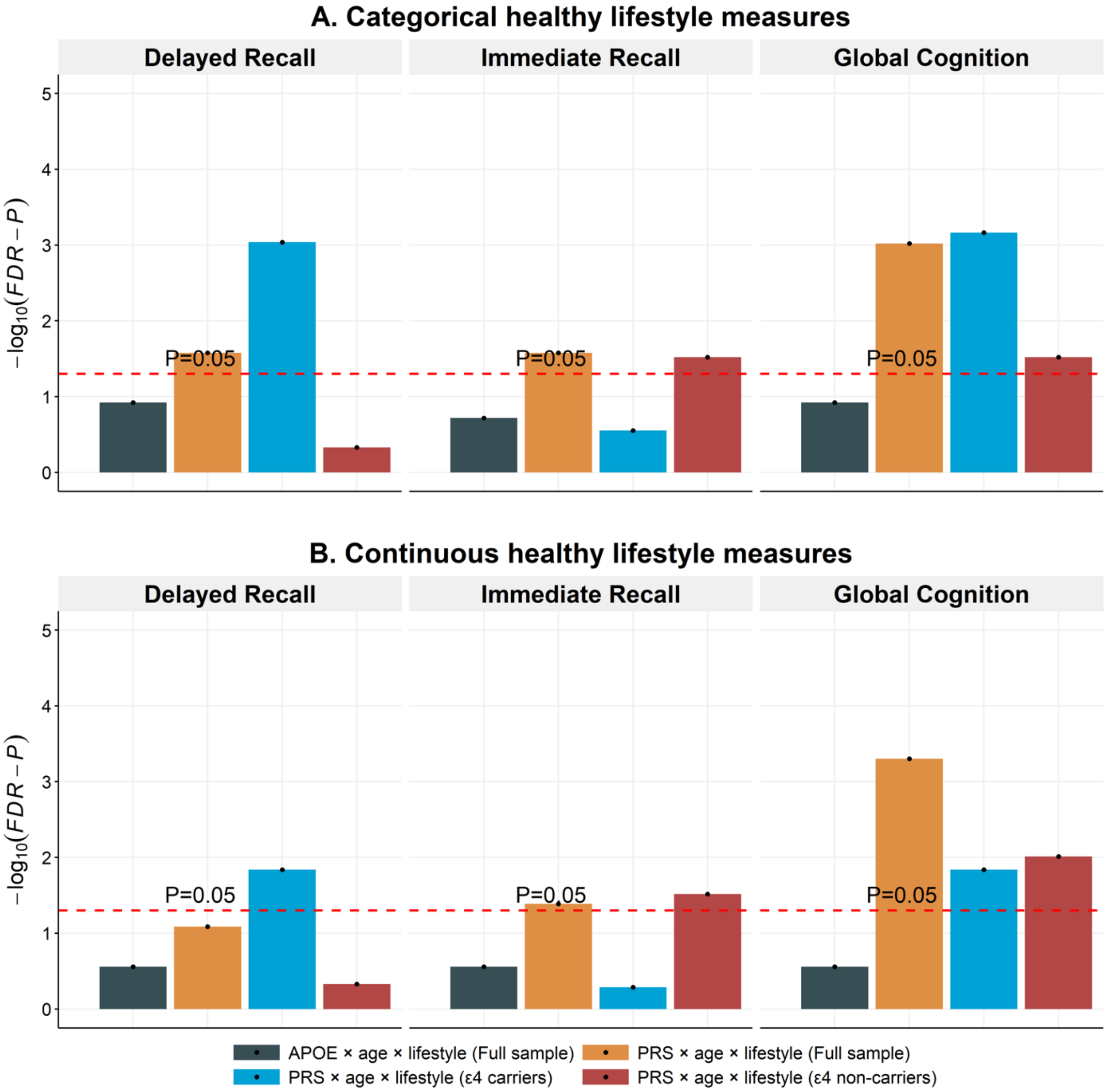

Supplementary Figure 5 presents the FDR corrected -log$_{10}$($P$) from the likelihood ratio tests for the three-way interaction terms between genetic risk predictor (*APOE* ε4 or PRS), adherence to healthy lifestyles (smoking, alchol consumption, physical exercises, and diet), and age in the HRS sample and sample stratified by *APOE* ε4 carrier status (for PRS analysis only). The likelihood ratio test statistic is calculated as the ratio between the log-likelihood of the nested model (model without three-way interaction terms) to the full model (model with genetic risk predictor*adherence to healthy lifestyles*polynomial age terms). All association tests were performed using linear mixed-effect model with random intercept at subject and household level.

Additional covariates include sex, education, practice effects, BMI, CESD-score, cohort, and the first five principal components of ancestry. For the *APOE* analyses in the full sample, we additionally adjusted PRS × Age interaction in all models to control for age heterogenous genetic effect caused by non-*APOE* genetic variants. For the PRS analyses in the full sample, we additionally adjusted *APOE* ε4 × Age interaction in all models to control for age heterogenous genetic effect caused by *APOE* genetic variants, as reported previously. Age is centered at year 65 and education is centered at the mean (13.7 years). Inverse probability weighting was adjusted to account for selection mortality in the HRS genetic sample. CESD-score = Center for Epidemiologic Studies Depression Scale score.

**Supplementary Figure 6. Simple slope estimates of PRS on domain specific- and global cognitive score for *APOE* ε4 carriers with a different number of healthy lifestyles (smoking, physical activity, diet, and alcohol consumption) and from age 55 to 85 in HRS (N = 447).**

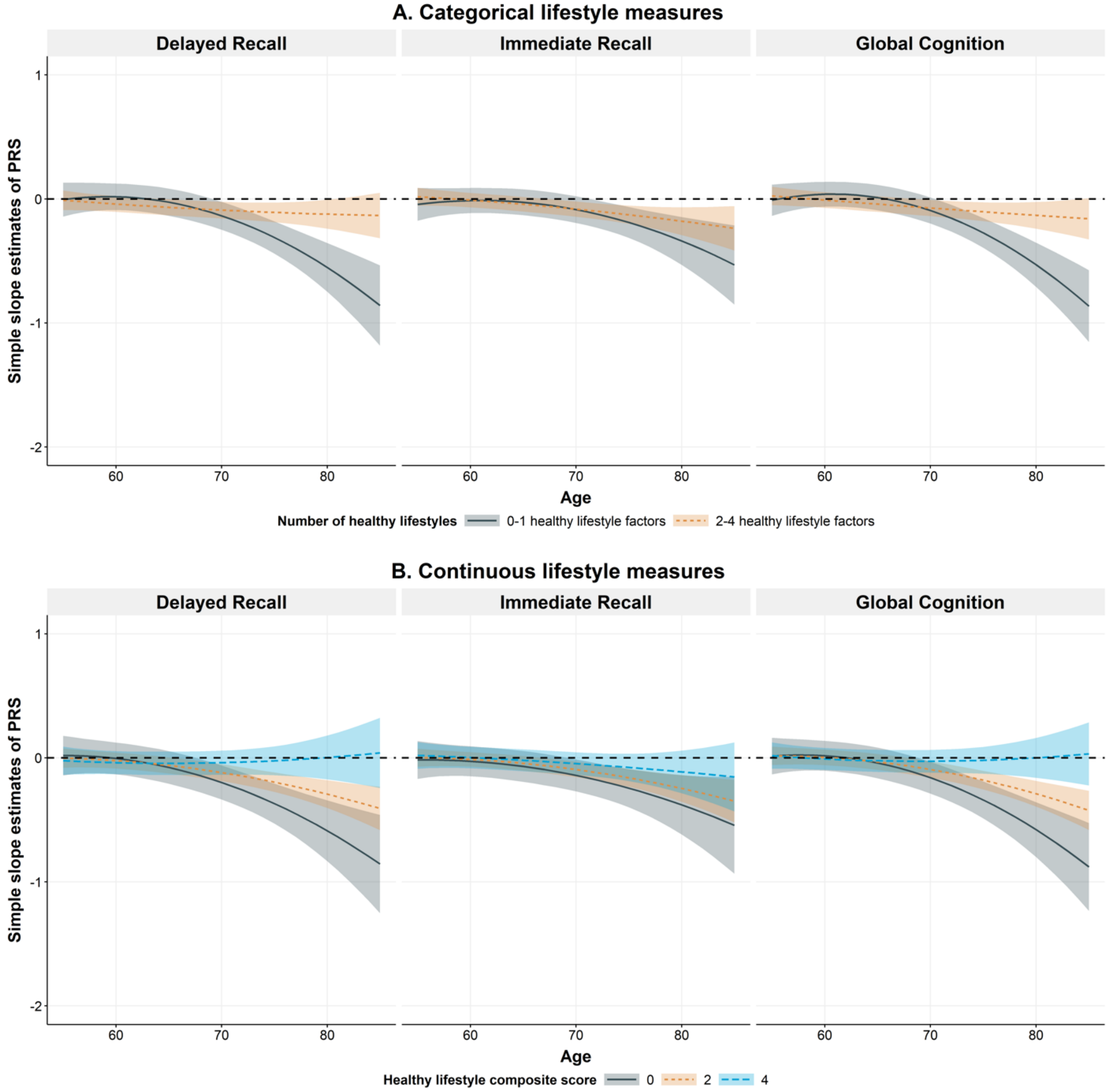

Supplementary Figure 6 shows the simple slope estimates of the PRS for *APOE* ε4 carriers with a different number of healthy lifestyles from age 55 to 85 on global and domain specific cognition score. For the categorical lifestyle measures, the grey and orange line represents the longitudinal trajectory of simple slope estimates of PRS among *APOE* ε4 carriers who have 0-1 healthy lifestyles, and 2-4 healthy lifestyles, respectively. For the continuous lifestyle measures, the grey, orange, and blue line represents the longitudinal trajectory of simple slope estimates of PRS among *APOE* ε4 carriers whose healthy lifestyle composite score is 0, 2, and 4, respectively. The simple slope estimates are calculated using the package “reghelper” in R and were based on the results which were obtained using the linear mixed-effect model and

adjusted for within-individual/household correlation. In addition to PRS, age (quadratic), adherence to healthy lifestyles, and their interactions, additional covariates include sex, education years, practice effect, cohort, BMI, CESD score, and the first five principal components of ancestry. Age is centered at year 65 and education is centered at the mean (13.7 years). Inverse probability weighting was adjusted to account for selection mortality in the HRS genetic sample. CESD-score = Center for Epidemiologic Studies Depression Scale score.

**Supplementary Figure 7. Simple slope estimates of PRS on domain specific- and global cognitive score for *APOE* ε4 non-carriers with different number of healthy lifestyles (smoking, physical activity, diet, and alcohol consumption) and from age 55 to 85 in HRS (N = 1,417).**

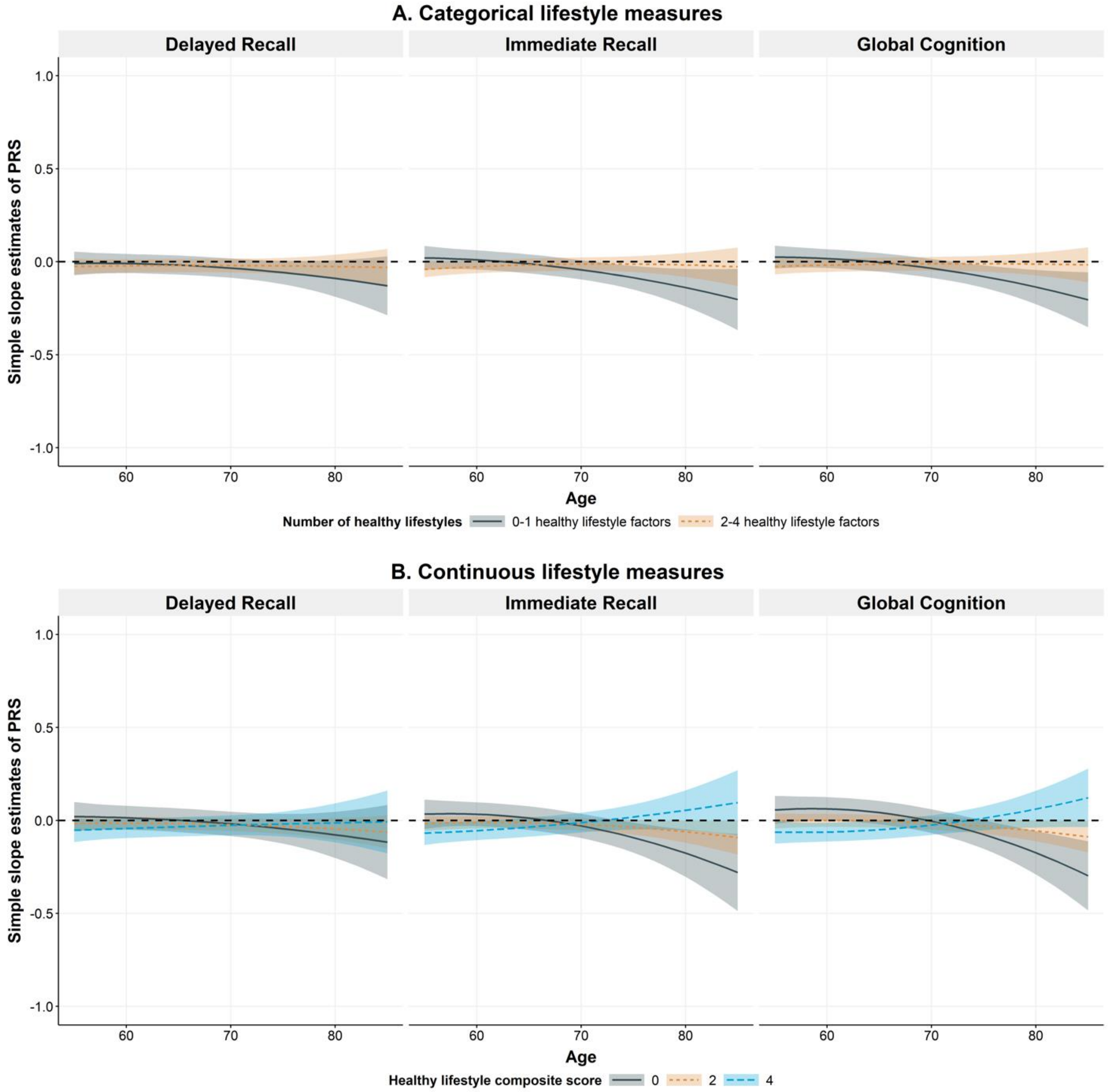

Supplementary Figure 7 shows the simple slope estimates of the PRS for *APOE* ε4 non-carriers with different number of healthy lifestyles from age 55 to 85 on global and domain specific cognition score. For the categorical lifestyle measures, the grey and orange line represents the longitudinal trajectory of simple slope estimates of PRS among *APOE* ε4 non-carriers who have 0-1 healthy lifestyles, and 2-4 healthy lifestyles, respectively. For the continuous lifestyle measures, the grey, orange, and blue line represents the longitudinal trajectory of simple slope estimates of PRS among *APOE* ε4 non-carriers whose healthy lifestyle composite score is 0, 2, and 4, respectively. The simple slope estimates are calculated using the package “reghelper” in R and were based on the results which were obtained using the linear mixed-effect model and

adjusted for within-individual/household correlation. In addition to PRS, age (quadratic), adherence to healthy lifestyles, and their interactions, additional covariates include sex, education years, practice effect, cohort, BMI, CESD score, and the first five principal components of ancestry. Age is centered at year 65 and education is centered at the mean (13.7 years). Inverse probability weighting was adjusted to account for selection mortality in the HRS genetic sample. CESD-score = Center for Epidemiologic Studies Depression Scale score.

**Supplementary Figure 8. Simple slope estimates of PRS on domain specific- and global cognitive score for *APOE* ε4 carriers with different number of healthy lifestyles (smoking, physical activity, and alcohol consumption) and from age 55 to 85 in HRS (N = 1,749).**

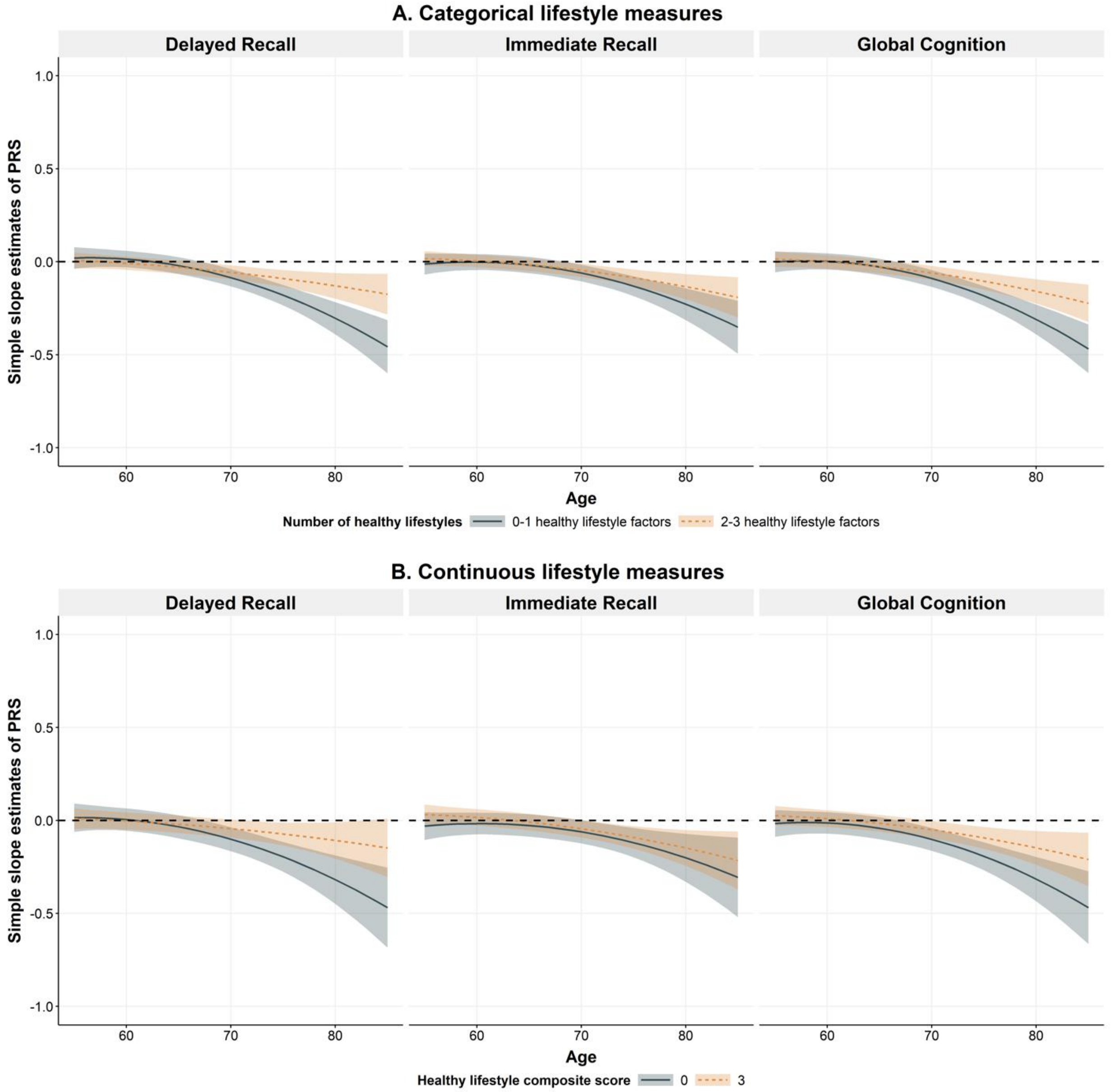

Supplementary Figure 8 shows the simple slope estimates of the PRS for *APOE* ε4 carriers with different number of healthy lifestyles from age 55 to 85 on global and domain specific cognition score. For the categorical lifestyle measures, the grey and orange line represents the longitudinal trajectory of simple slope estimates of PRS among *APOE* ε4 non-carriers who have 0-1 healthy lifestyles, and 2-3 healthy lifestyles, respectively. For the continuous lifestyle measures, the grey and orange line represents the longitudinal trajectory of simple slope estimates of PRS among *APOE* ε4 carriers whose healthy lifestyle composite score is 0, and 3, respectively. The simple slope estimates are calculated using the package “reghelper” in R and were based on the results which were obtained using the linear mixed-effect model and

adjusted for within-individual/household correlation. In addition to PRS, age (quadratic), adherence to healthy lifestyles, and their interactions, additional covariates include sex, education years, practice effect, cohort, BMI, CESD score, and the first five principal components of ancestry. Age is centered at year 65 and education is centered at the mean (13.5 years). Inverse probability weighting was adjusted to account for selection mortality in the HRS genetic sample. CESD-score = Center for Epidemiologic Studies Depression Scale score.

**Supplementary Figure 9. Simple slope estimates of PRS on domain specific- and global cognitive score for *APOE* ε4 non-carriers with different number of healthy lifestyles (smoking, physical activity, and alcohol consumption) and from age 55 to 85 in HRS (N = 4,937).**

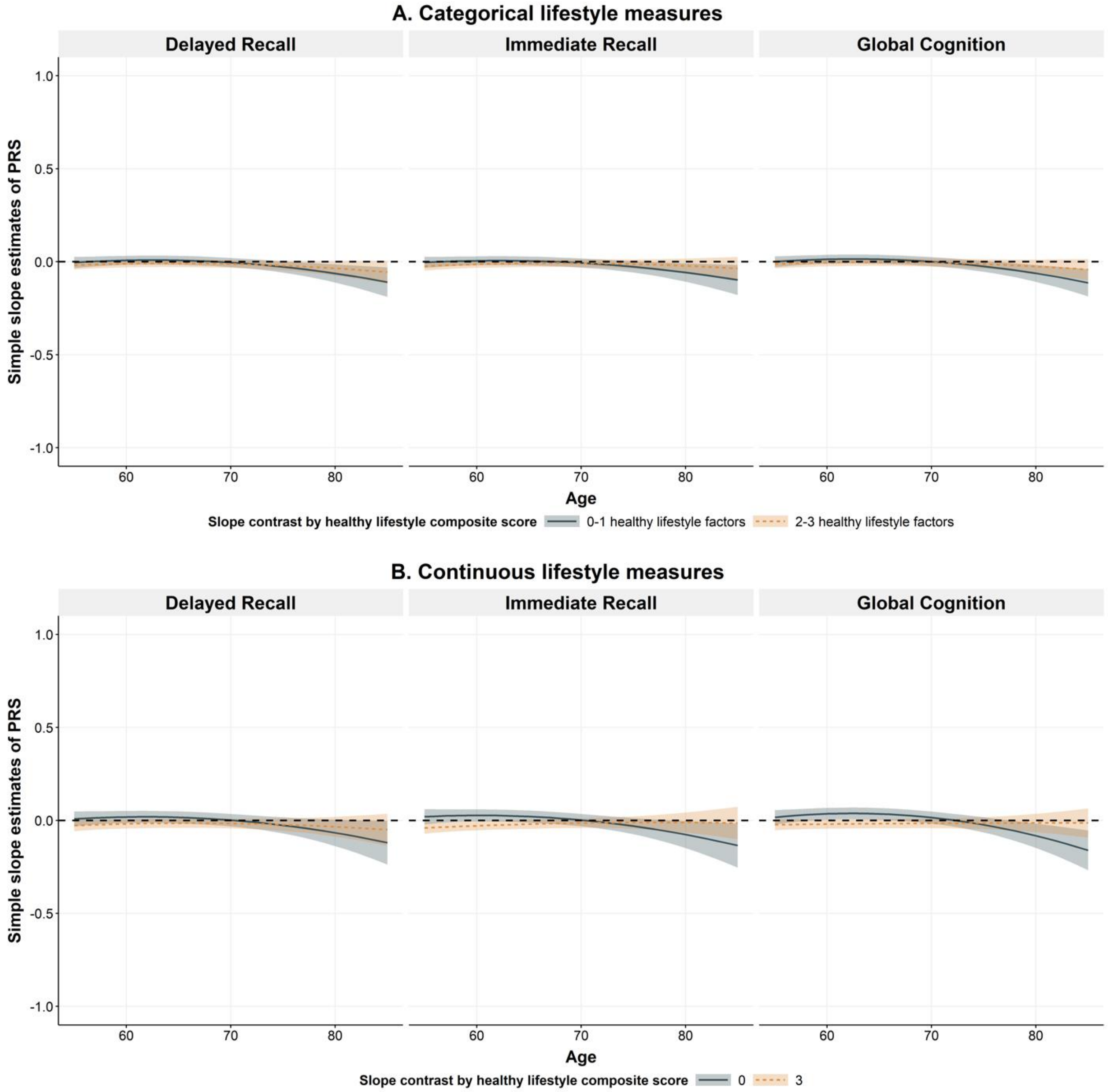

Supplementary Figure 9 shows the simple slope estimates of the PRS for *APOE* ε4 non-carriers with different number of healthy lifestyles from age 55 to 85 on global and domain specific cognition score. For the categorical lifestyle measures, the grey and orange line represents the longitudinal trajectory of simple slope estimates of PRS among *APOE* ε4 non-carriers who have 0-1 healthy lifestyles, and 2-3 healthy lifestyles, respectively. For the continuous lifestyle measures, the grey, and orange line represents the longitudinal trajectory of simple slope estimates of PRS among *APOE* ε4 non-carriers whose healthy lifestyle composite score is 0, and 3, respectively. The simple slope estimates are calculated using the package “reghelper” in R and were based on the results which were obtained using the linear mixed-effect model and

adjusted for within-individual/household correlation. In addition to PRS, age (quadratic), adherence to healthy lifestyles, and their interactions, additional covariates include sex, education years, practice effect, cohort, BMI, CESD score, and the first five principal components of ancestry. Age is centered at year 65 and education is centered at the mean (13.5 years). Inverse probability weighting was adjusted to account for selection mortality in the HRS genetic sample. CESD-score = Center for Epidemiologic Studies Depression Scale score.

2. CDC. National Health Interview Survey - Adult Alcohol Use Information. https://www.cdc.gov/nchs/nhis/alcohol/alcohol_glossary.htm

10. Health and Retirement Study, (Health and Retirement Study (HRS) Genotype Data) restricted dataset. Produced and distributed by the University of Michigan with funding from the National Institute on Aging (grant number NIA U01AG009740). Ann Arbor, MI,. Published online 2022.
